# Immune evolution from preneoplasia to invasive lung adenocarcinomas and underlying molecular features

**DOI:** 10.1101/2020.07.11.20142992

**Authors:** Hitoshi Dejima, Xin Hu, Runzhe Chen, Jiexin Zhang, Junya Fujimoto, Edwin R. Parra, Cara Haymaker, Shawna Hubert, Dzifa Duose, Luisa M. Solis, Dan Su, Junya Fukuoka, Kazuhiro Tabata, Hoa Pharm, Nicholas Mcgranahan, Baili Zhang, Jie Ye, Lisha Ying, Latasha Little, Curtis Gumbs, Chi-Wan Chow, Marcos Roberto Estecio, Myrna C.B. Godoy, Mara B. Antonoff, Boris Sepesi, Harvey Pass, Carmen Behrens, Jianhua Zhang, Ara A. Vaporciyan, John V. Heymach, Paul Scheet, J. Jack Lee, P. Andrew Futreal, Alexandre Reuben, Humam Kadara, Ignacio Wistuba, Jianjun Zhang

## Abstract

How anti-cancer immunity shapes early carcinogenesis of lung adenocarcinoma (ADC) is unknown. We characterized immune contexture of invasive lung ADC and its precursors by transcriptomic immune profiling, T cell receptor (TCR) sequencing and multiplex immunofluorescence. Our results demonstrated that anti-tumor immunity evolved as a continuum from lung preneoplasia, to preinvasive ADC, minimally-invasive ADC and frankly invasive lung ADC with a gradually less effective and more intensely regulated immune response including down-regulation of immune-activation pathways, up-regulation of immunosuppressive pathways, higher infiltration of CD4+ T cells, lower infiltration of CD8+ T cells, decreased T cell clonality, and lower frequencies of top T cell clones in later stages. Driver mutations, HLA loss, chromosomal copy number aberrations and DNA methylation changes may collectively impinge host immune responses and facilitate immune evasion as a potential mechanism underlying outgrowth of the most fit subclones in preneoplasia into dominant clones in invasive ADC.

**SIGNIFICANCE:** There has been a drastic increase in the detection of lung nodules, many of which are lung ADC precursors. The management of these lung nodules is controversial. We discovered that immune activation and evasion have started at preneoplastic stage and lung ADC precursors may exhibit an overall better-preserved anti-tumor immune contexture suggesting therapeutic strategies reprograming the immune microenvironment in patients with lung ADC precursors prior to further immunosuppression in invasive lung cancers may be beneficial. These findings have served as the critical scientific rationale for our immunoprevention clinical trial IMPRINT-Lung (NCT03634241) recruiting individuals diagnosed with lung nodules at high risk developing invasive lung cancers.

## INTRODUCTION

Despite significant advances in its management, lung cancer remains the leading cause of cancer death worldwide (Siegel et al., 2020; Tan et al., 2016), largely due to late diagnosis at advanced stages when cures are generally unachievable (Barnett, 2017; Borghaei et al., 2015; Siegel et al., 2020). Computed tomography (CT) scan-guided lung cancer screening has demonstrated a reduction of lung cancer mortality by 26%-61% (de Koning et al., 2020) highlighting the importance of early detection and intervention. These findings suggest that early cancer interception is crucial to reduce lung cancer incidence and mortality. Yet, to date, randomized clinical trials on primary lung cancer prevention have only produced disappointing results (de Koning et al., 2020), primarily due to our rudimentary knowledge of early phases in lung cancer development. Improved understanding of targetable molecular mechanisms underlying early lung carcinogenesis may accelerate the development of precise diagnostic as well as effective preventive and therapeutic strategies.

Lung adenocarcinoma (ADC) is the most common histological subtype of lung cancer. Recent studies has postulated that lung ADC may arise from atypical adenomatous hyperplasia (AAH), the only recognized preneoplasia to lung ADC (Aoyagi et al., 2001; Chiosea et al., 2007; Kitamura et al., 1999; Maeshima et al., 2010; Min et al., 2010; Noguchi, 2010; Seki and Akasaka, 2007), which evolves into preinvasive adenocarcinoma *in situ* (AIS) (Weichert and Warth, 2014), to micro-invasive lesion termed minimally invasive adenocarcinoma (MIA) (Aoyagi et al., 2001; Travis et al., 2011) and eventually frankly invasive ADC (Aoyagi et al., 2001; Travis et al., 2011). Early-stage lung ADCs and their precursors usually present as lung nodules with distinct radiologic features called ground glass opacity (GGO). These lung nodules are often referred as indeterminate pulmonary nodules (IPN) without histologic diagnosis as the diagnostic yield from biopsy of GGO-predominate nodules is low and surgery is not the standard of care. This has subsequently led to the scarcity of appropriate materials to study the molecular profiles of lung ADC precursors (Izumchenko et al., 2015).

Carcinogenesis results from progressive accumulation of molecular abnormalities (molecular evolution) (Vogelstein et al., 2013) and escape from host immune surveillance (immunoediting) (Schreiber et al., 2011). Our recent gene expression and genomic pilot studies on lung ADC precursors have demonstrated distinct transcriptomic features (Sivakumar et al., 2017) and progressive genomic evolution along the spectrum of AAH to AIS, MIA and ADC (Hu et al., 2019). However, the extent to which immunoediting sculpts early carcinogenesis of lung ADC and the underlying genomic and epigenetic alterations associated with these immune features still remain to be determined. In the current study, we performed immune gene expression profiling, T cell receptor (TCR) sequencing and multiplex immunofluorescence (mIF) staining on a cohort of resected AAH, AIS, MIA and invasive ADC lesions and paired morphologically normal lung tissues (NL) to delineate the evolution of immune contexture, particularly T cell landscape across different stages of early lung ADC pathogenesis. We further leveraged whole exome sequencing (WES) (Hu et al., 2019) and methylation data (Hu et al., 2020) from the same cohort of IPNs to underscore the genomic and epigenetic alterations that may impinge on these immune features (**Table S1** and **Figure 1**).

**Figure 1.**
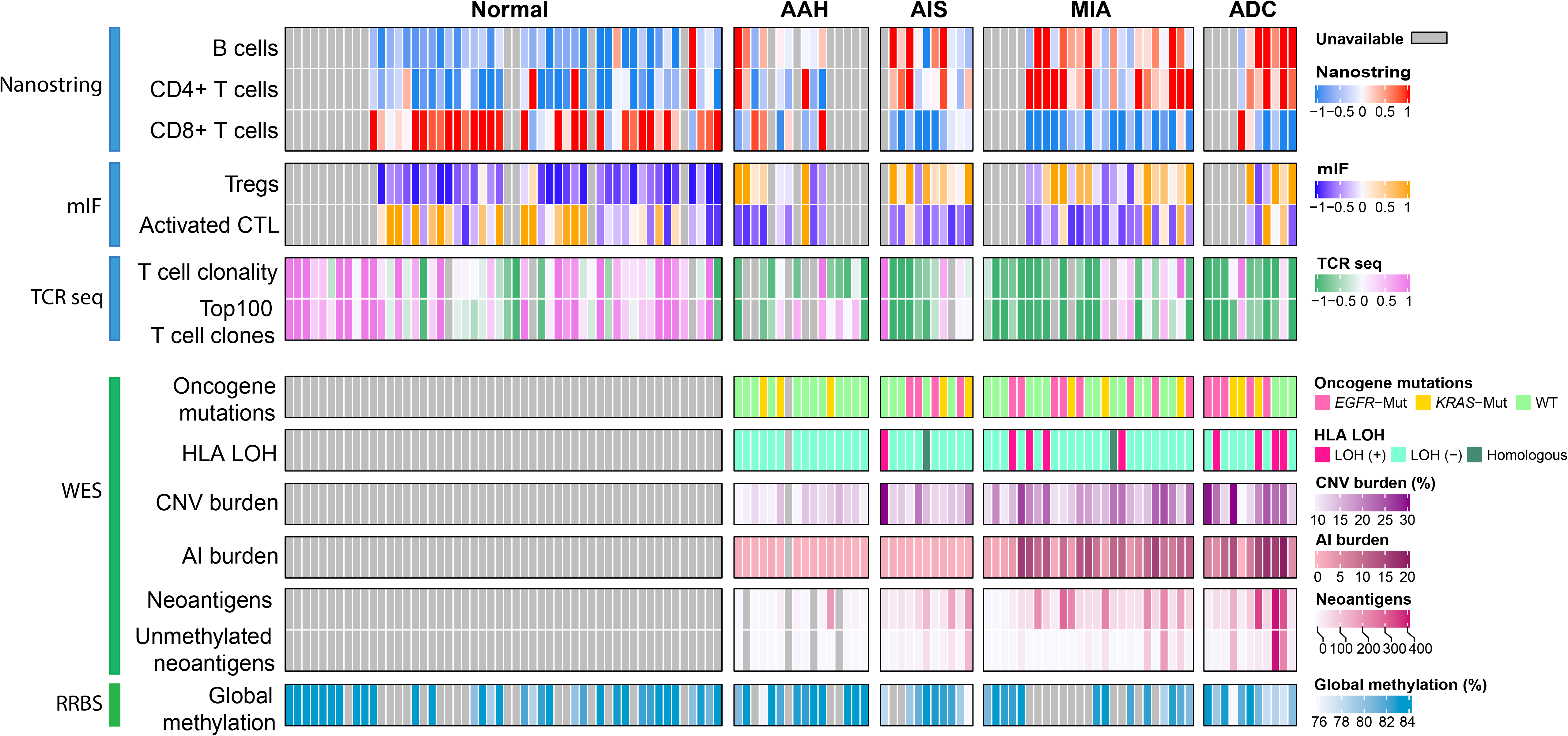
The immune evolution from preneoplasia to invasive lung adenocarcinoma and associated genomic and epigenomic features. Infiltration of B cells, CD4+ T cells, CD8+ T cells inferred from immune gene expression using TIMER; regulatory T cells (Treg) and activated cytotoxic T lymphocytes (CTL) measured by multiplex immunofluorescence (mIF); T cell clonality and frequency of the top 100 T cell clones by TCR sequencing are shown in upper panel. Genomic alterations from whole exome sequencing (WES) including *EGFR/KRAS* mutations, HLA loss, copy number variation (CNV) burden, allelic imbalance (AI) burden, total number of mutations associated with predicted neoantigens, total number of mutations associated with predicted neoantigens without promoter methylation; global methylation status accessed by reduced representation bisulfite sequencing (RRBS) are shown in bottom panel. AAH: typical adenomatous hyperplasia. AIS: adenocarcinoma in situ. MIA: minimally invasive adenocarcinoma. ADC: invasive adenocarcinoma.

## RESULTS

### Progressive decrease in overall immunity mirrors evolution from preneoplasia to invasive lung adenocarcinoma

To assess dynamic changes in the immune contexture during early lung carcinogenesis, we performed immune profiling using the nCounter PanCancer Immune Profiling Panel (NanoString), which includes 770 genes from 14 different immune cell types, common checkpoint inhibitors, cancer/testis antigens, and genes covering both the adaptive and innate immune response (Chen et al., 2016) on 47 resected pulmonary nodules (n=9 for AAH, n=11 for AIS, n=21 for MIA, and n=6 for invasive ADC) and paired NL tissues (n=38). There were no differences in age (p=0.55, Kruskal-Wallis test), sex (p=0.31, fisher’s exact test) or smoking status (p=0.35, fisher’s exact test) between patients with pulmonary nodules of different histologic stages. In total, 291 genes were differentially-expressed (DEGs). Interestingly, changes in the majority of DEGs, regardless of their direction, exhibited a progressive pattern along the spectrum from NL, to AAH, AIS, MIA and ADC (**Table S2**). Examples of progressively increased genes included immune suppressive genes *CD47* (protection of cancer cells from immune cell killing) (Soto-Pantoja et al., 2014), *CD276* (inhibition of immune responses) (Picarda et al., 2016) and *CTLA4* (checkpoint molecule) (Pardoll, 2012), while progressively decreased genes included *ENTPD1* (expressed on tumor-specific T cells) (Bastid et al., 2013), granzyme B *(GZMB)* and perforin 1 *(PRF1)* (two cytotoxic molecules produced by T lymphocytes and natural killer cells (NK cells) (Prakash et al., 2014; Tschopp et al., 1986) (**Figure S1**). Functional pathway analysis of DEGs revealed 26 significantly de-regulated pathways associated with neoplastic evolution from NL to invasive ADC, of which, 23 were down-regulated (**Figure 2A**). On the other hand, all three up-regulated pathways (systemic lupus erythematosus (SLE) in B cell signaling, T cell exhaustion signaling and PARP signaling pathways) could potentially impair immune response (Pan et al., 2020; Pantelidou et al., 2019; Wherry and Kurachi, 2015). These results indicated an overall decreased immunity in later-stage lesions.

**Figure 2.**
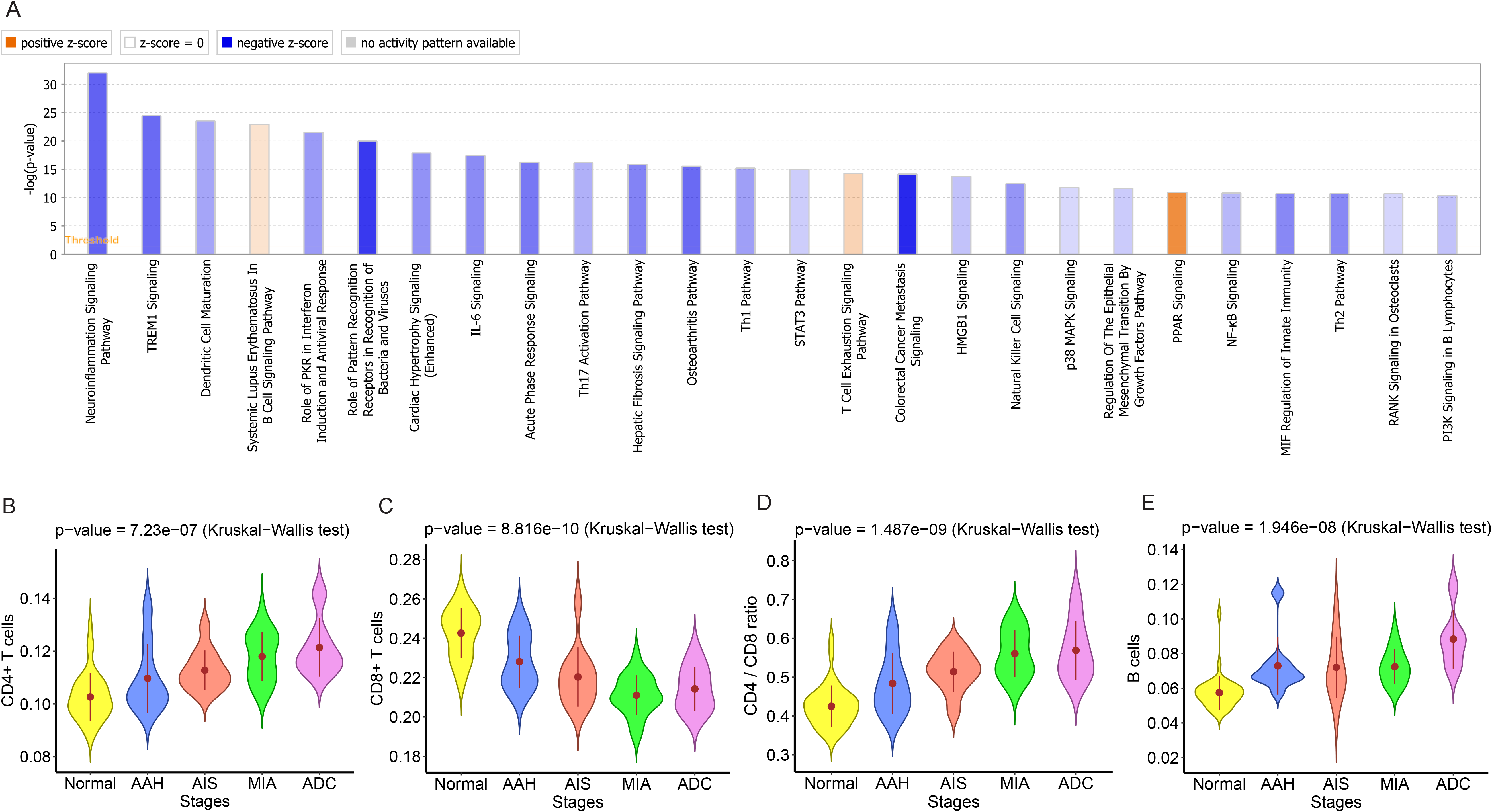
Progressively changes of immune cell infiltrations from preneoplasia to invasive lung adenocarcinoma. (**A**) Significantly enriched functional pathways based on the 291 differentially expressed genes by Ingenuity pathway analysis (IPA®; Ingenuity Systems) software. Pathways with –log (p-value) > 10 (p-values are obtained from Fisher’s right-tailed exact test) and an absolute z-score > 0.5 are shown. Pathways that were predicted to be inhibited (negative Z scores) in later stages are in blue and pathways that were predicted to be activated (positive Z scores) in later stages are in orange. The heights of the bars indicate the significance of the enrichment (-log (p-value)) and the scales of the orange or blue colors represent the predicted directionality. Fractions of immune cells including CD4+ T cells (**B**), CD8+ T cells (**C**), CD4/CD8 ratio (**D**) and B cells (**E**) were estimated using TIMER based on the gene expression using nCounter PanCancer Immune Profiling Panel. Error bars indicate 95% confidence intervals and solid point represent mean value in each stage. The difference of cell fraction among different stages was evaluated using the Kruskal-Wallis H test. NL: Normal lung tissue, AAH: Atypical adenomatous hyperplasia, AIS: Adenocarcinoma in situ, MIA: Minimally invasive adenocarcinoma, ADC: Invasive adenocarcinoma.

We next de-convoluted gene expression profiling data using TIMER (Li et al., 2020) to evaluate changes in immune cell composition. As shown in **Figure 2B**, CD4+ T lymphocyte infiltration progressively increased from NL to invasive ADC. Conversely, infiltration of CD8+ T lymphocytes progressively decreased with neoplastic evolution (**Figure 2C**) leading to significantly higher CD4/CD8 ratio in later-stage lesions (**Figure 2D**). Additionally, B cell infiltration progressively increased along the spectrum from NL to invasive ADC (**Figure 2E**). Of particular interest, all 6 tertiary lymphoid structure (TLS) markers CD19, MS4A1 (CD20), CXCL13, CXCR5, CCR7 and CCL19 (Cabrita et al., 2020) (**Figure S2**) also progressively increased from NL to invasive ADC indicating a progressive TLS aggregation, which may play a role in follicular regulatory T cell-mediated CD8+ T cell exclusion (Wang et al., 2020). Indeed, these TLS markers were positively associated with CD4+ T cell infiltration, but negatively associated with CD8+ T cell infiltration (**Figure S3**). To validate these findings, we applied TIMER to RNAseq data from an independent cohort previously published by our group and observed similar results (**Figure S4**) (Sivakumar et al., 2017).

Taken together, these results suggest that immune evolution progressed as a continuum from preneoplasia to invasive lung ADC with a gradually less effective and more intensely regulated immune response.

### Dynamic changes in T cell phenotype and infiltration define histologic stages of early ADC development

We next performed multiplex immunofluorescence (mIF) using antibodies against cytokeratin (CK), CD3, CD8, PD-1, PD-L1, CD68, CD45RO, GZMB and FoxP3 (**Figure 3A, Figure S5** and **Figure S6**) on a subset of pulmonary nodules (n=9 for AAH, n=10 for AIS, n=21 for MIA, and n=6 for invasive ADC) and paired NL (n =7) to assess the dynamic changes of various subtypes of T cells and their interaction with (pre-)malignant cells during early lung carcinogenesis. Densities of infiltrating activated CTLs (CD3+CD8+GZMB+) and regulatory T cells (Treg, CD3+CD8-FoxP3+) assessed by mIF were positively correlated with corresponding CD8+ and CD4+ T cell infiltrate levels derived from gene expression profiling, respectively (**Figure S7A-B**). Furthermore, Treg/activated CTL ratio closely recapitulated CD4/CD8 ratio inferred from immune gene expression profiling (**Figure S7C**). Correlations among immune subsets and between immune components and epithelial cells across IPNs of different histologic stages demonstrated that in normal tissues, densities of CK+ and CK+PD-L1+ cells were positively correlated, as were Tregs and memory T cells, cytotoxic and activated cytotoxic T cells (**Figure S8A**). For AAH, only total macrophages and activated cytotoxic T cells were positively correlated (**Figure 3B**). In AIS, PD-L1+ macrophages were positively correlated with Tregs, antigen-experienced T cells and cytotoxic T cells (**Figure 3C**). For MIA, CK cells were negatively correlated with total macrophages and memory T cells, while Tregs were positively correlated with antigen-experienced T cells and memory T cells were correlated with cytotoxic and activated cytotoxic T cells (**Figure 3D**). Finally, in ADC, PD-L1+CK+ cells were correlated with total and PD-L1+ macrophages, PD-L1 + macrophages were correlated with total macrophages, and Tregs were correlated with memory T cells (**Figure S8B**). Overall, relationships between cell types varied widely across stages, highlighting the dynamic nature of tumor-immune interactions during tumor evolution.

**Figure 3.**
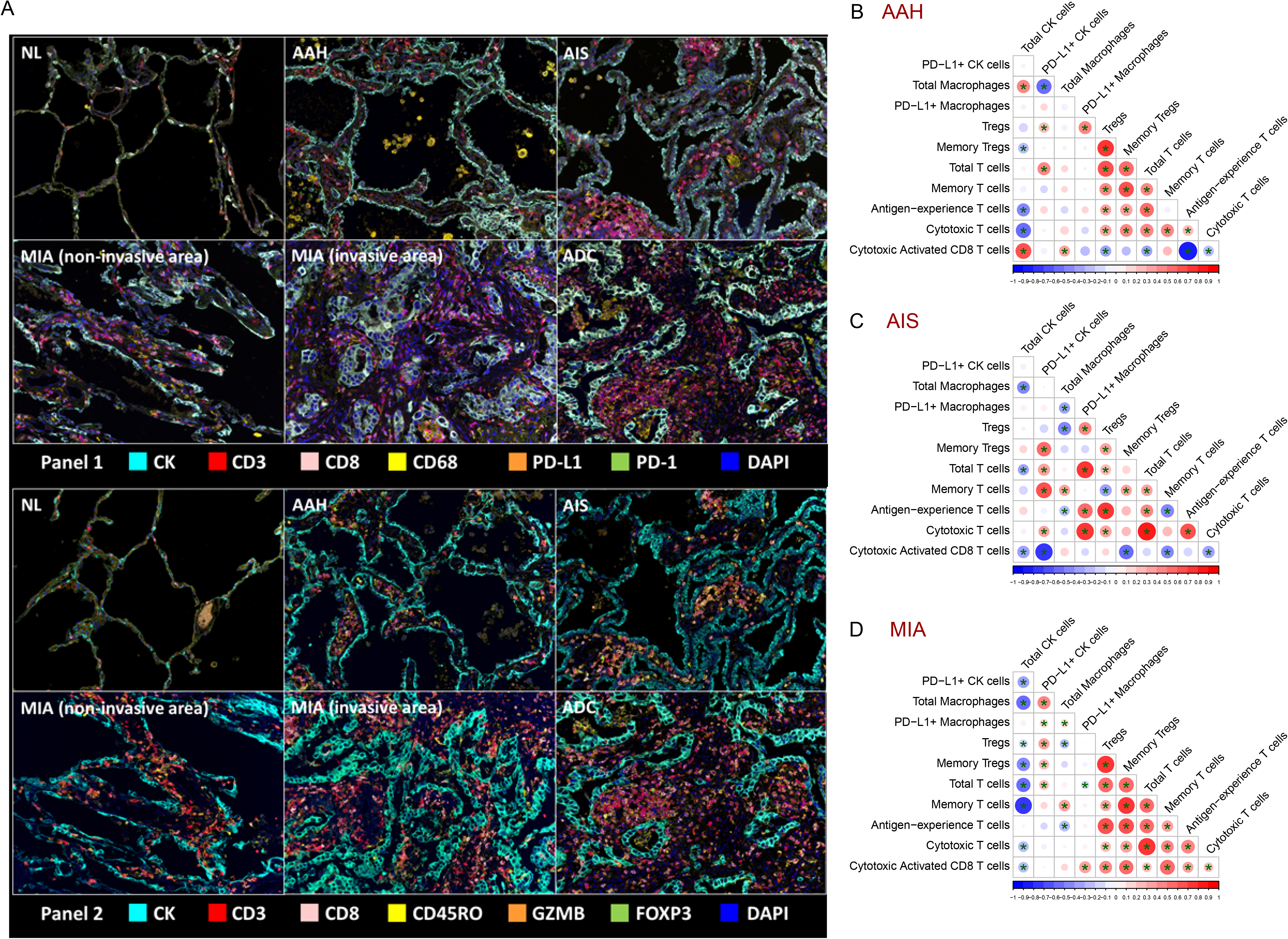
Dynamic changes of various immune cells and their interaction with epithelial cells across preneoplasia to invasive lung adenocarcinoma. (**A**) Representative multiplex immunofluorescence (mIF) images each histologic stage analyzed by panel 1 and panel 2 markers. The correlation between immune cell subtypes and CK+ epithelial cells measured by mIF in AAH (**B**), AIS (**C**) and MIA (**D**). Significant correlation was marked with ^*^ (p<0.05). Red circles indicate positive correlations and blue circles indicate negative correlations. The size of circles is proportional to the spearman’s correlation co-efficient between each pair of cells. AAH: Atypical adenomatous hyperplasia, AIS: Adenocarcinoma in situ, MIA: Minimally invasive adenocarcinoma.

### Progressively divergent TCR repertoire with neoplastic progression

Because of the central role of T cells in tumor surveillance, we next sought to investigate the T cell repertoire (Shah et al., 2011) by multiregional T cell receptor (TCR) sequencing on 13 AAH, 11 AIS, 23 MIA and 10 ADC as well as 49 NL. We first assessed the T cell diversity using Inverse Simpson Index (Kaplinsky and Arnaout, 2016). T cell diversity was positively correlated with infiltration of CD4+ T cells derived from gene expression profiling (**Figure S9A**) as well as densities of Tregs from mIF (**Figure S9B**) and increased steadily from AAH to AIS, MIA and ADC (p<0.0001) **(Figure 4A)** in line with higher CD4+ T cells in later stage lesions **(Figure 2B)**. We then compared the distributions of T cell clonality across histologic stages, focusing on the most expanded T cell clones (Mansfield et al., 2018; Wang et al., 2019). As shown in **Figure 4B** and **Figure S10**, the top 10, 100, 200, and 500 clones accounted for a higher frequency in the NLs, and gradually decreased in AAH, AIS, MIA and invasive ADC (p<0.0001) implying suppressed T cell expansion during early carcinogenesis of lung ADC.

**Figure 4.**
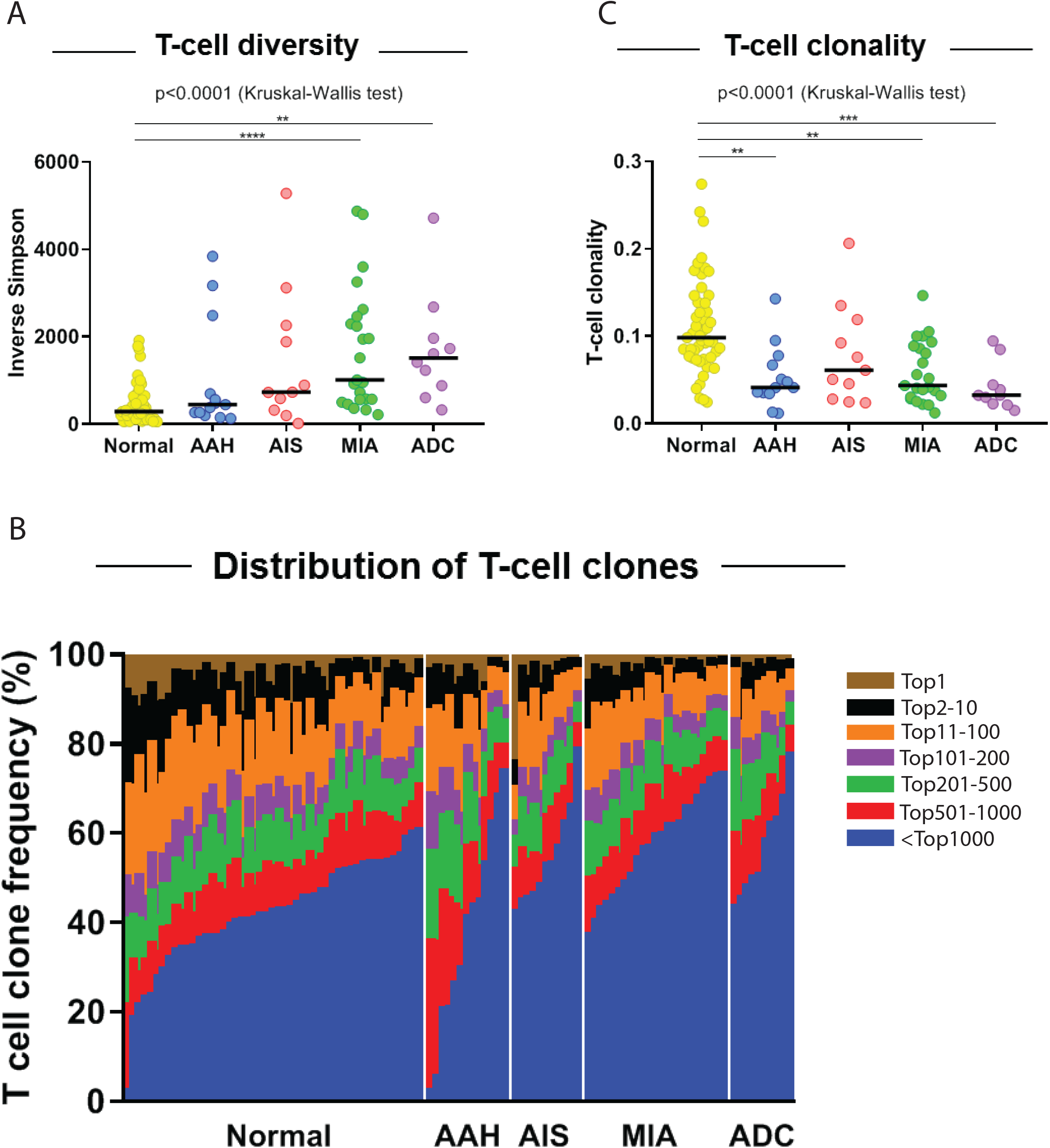
Dynamic change in T cell repertoire from preneoplasia to invasive lung adenocarcinoma. T cell diversity (**A**) and T cell clonality (**C**) in normal lung, AAH, AIS, MIA and invasive ADC. (**B**) Distribution of T cell clones with frequency of top 1 (brown), top 2 to 10 (black), top 11 to 100 (orange), top 101 to 200 (purple), top 201 to 500 (green), top 501 to 1000 (red) and beyond 1000 (blue) in normal lung, AAH, AIS, MIA and invasive ADC lesions.

### Suppression of the T cell repertoire may occur at the preneoplastic stage

To further understand the T cell response at different histologic stages, we calculated T cell clonality, a metric indicating expansion and activation of T cell clones. T cell clonality was positively correlated with infiltration of CD8+ T cells (r=0.561, p=4.863e-06), activated CTLs (r=0.318, p=7.340e-03), *GZMB* expression (r=0.319, p=5.068e-02) and Th1 cytokines (r=0.473, p=3.923e-05) (**Figure S11A-D**), while negatively correlated with infiltration of CD4+ T cells (r=-0.385, p=8.993e-04) and Tregs (r=-0.477, p=2.965e-05) (**Figure S11E-F**) suggesting that T cell clonality was mainly driven by clonal expansion of activated CTLs, with Tregs inhibiting CTL responses. Comparing T cell clonality among different stages revealed the highest clonality in NL **(Figure 4C)** consistent with our previous findings (Reuben et al., 2020). T cell clonality declined from NL to AAH increasing in AIS/MIA and declining to its lowest level in ADC **(Figure 4C)**. These results suggest that early ADC pathogenesis is associated with local immunosuppression in TCR repertoire that may have commenced at the preneoplastic stage.

### Driver mutations affect the immune response in pre/early ADC

We next sought to explore the molecular features associated with the immune contexture observed in these lesions. We first explored the two most frequently mutated driver genes in this cohort *EGFR* and *KRAS* (Hu et al., 2019). Compared to EGFR-mutant lesions or double wild-type lesions, *KRAS*-mutant lesions exhibited highest CD8+ T cell infiltration and lowest CD4/CD8 T cell ratio inferred from immune gene expression; highest CTL infiltration, highest effector memory cytotoxic T cell infiltration, highest CTL/Treg ratio from mIF as well as highest T cell clonality measured by TCR sequencing **(Figure S12)**. These trends remained similar when analyzing each histologic stage (**Figure S13**). These findings are consistent with our previous studies in invasive lung cancers (Reuben et al., 2020) and emphasize the interplay between oncogene mutations and immune surveillance during early pathogenesis of lung adenocarcinoma.

### Chromosomal copy number variations and HLA loss may contribute to impaired T cell responses

In light of recent studies suggesting immune evasion could be facilitated by an inability to present neoantigens due to loss of HLA (McGranahan et al., 2017), we applied the LOHHLA (loss of heterozygosity in HLA) algorithm (McGranahan et al., 2017) to WES data from these lesions (Hu et al., 2019). LOH at the HLA loci was observed in 7%, 15% and 33% of AIS, MIA and ADC respectively, but in none of AAH lesions **(Figure S14A**, p=0.005, Chi-Square test). Correlation with T cell infiltration derived from immune gene expression profiling demonstrated that lesions with HLA LOH had similar CD4+ T cell infiltration, but trends of lower CD8+ T cell infiltration and higher CD4/CD8 ratio although the difference did not reach statistical significance **(Figure S14B-D**).

Chromosomal copy number variation (CNV) has been reported to impact immune microenvironment across different cancers (Davoli et al., 2017). We next investigated whether CNV affected the immune landscape of these lung ADC precursors. Correlation with T cell infiltration derived from gene expression profiling demonstrated burden of allelic imbalance (AI), a subtle form of CNV, was negatively correlated with CD8+ T cell infiltration, but positively correlated with CD4+ T cell infiltration and CD4/CD8 T cell ratio **(Figure S15A-C**). Similar trend was also observed with CNV burden, although these differences did not reach statistical significance **(Figure S15D-F**). Interestingly, HLA-LOH positive lesions exhibited significantly higher AI burden and CNV burden compared to that of HLA-LOH negative lesions **(Figure S16**). One plausible explanation is that the development AI, CNV or HLA LOH resulted from chromosomal instability (CIN) and cell clones with CIN may lead to higher CNV/AI burdens as well as increased likelihood of HLA loss, which could subsequently enable these cells escaping from anti-tumor immune surveillance and developing into dominant clones in the later-stage of neoplastic lesions.

### Methylation status interacts with genomic alterations and impacts the immune response during early lung cancer pathogenesis

Somatic mutations play central roles in activating anti-tumor immune responses through generating neoantigens that can be recognized by T cells (McGranahan et al., 2016; Yarchoan et al., 2017). As shown in **Figure S17**, a progressive increase in total mutation burden was observed from AAH to AIS, MIA and ADC. However, analysis of methylation data from reduced representation bisulfite sequencing (RRBS) of the same cohort of IPNs (Hu et al., 2020) revealed that a significantly larger proportion of genes exhibited promoter hypermethylation (>30% CpG methylated) in later-stage lesions, thus potentially dampening the expression of neoantigens in later stage lesions. These data suggest that promoter hypermethylation could contribute to neoantigen depletion and immune escape. Furthermore, we assessed the impact of global methylation status, using long interspersed transposable elements-1 (LINE-1) as a surrogate marker (Kankava et al., 2018; Ohka et al., 2011; Saito et al., 2010), on the immune microenvironment in these lesions. Overall, global methylation level was negatively associated with CD4+ T cell infiltration and CD4/CD8 ratio derived from gene expression profiling, Tregs infiltration and Treg/CD8 ratio from mIF (**Figure S18**) suggesting decreased global methylation was associated suppressive immune contexture.

## DISCUSSION

Cancer evolution is shaped by the interaction between cancer cells and host immune surveillance, a process termed immunoediting consisting of elimination, equilibrium, and escape phases (Dunn et al., 2002; Dunn et al., 2004a; Dunn et al., 2004b). It is well documented that the majority of human cancers are infiltrated with various immune cells, but often in an immunosuppressive microenvironment as remnant evidence of immunoediting (Mittal et al., 2014; O’Donnell et al., 2019). T cell immunity is significantly compromised even in stage I lung cancers (Lavin et al.; Reuben et al., 2020) indicating that these cancers have already begun evading immune surveillance. However, when and how the elimination and equilibrium phases occur over the lung cancer evolution continuum is unknown. Therefore, investigating the molecular and immune landscape of lung premalignancies is warranted to elucidate the timing of immune activation/evasion and its underlying molecular mechanisms during early lung carcinogenesis.

In the current study, we leveraged a relatively large cohort of resected lung ADC precursors with available genomic and epigenetic profiling data to characterize the dynamic changes in the immune contexture across different consecutive developmental stages by gene expression profiling, mIF and TCR sequencing. To the best of our knowledge, this is the first systemic and comprehensive study on immune evolution during early carcinogenesis of lung ADC and its potential underlying molecular basis. Overall, there was a more suppressive and tightly controlled immune response, particularly T cell response, in later stage lesions highlighting that the dynamic interaction between cancer cells and host immune surveillance was overall evolving toward immune escape along with pathogenesis of lung ADC. Notably, dynamic changes in immune contexture progressed as a continuum from preneoplasia AAH to pre-invasive AIS, to micro-invasive MIA and finally frankly invasive ADC without obvious “step-wise” major leaps at transitions between different histologic stages. This is consistent with the overall progressive genomic evolution from AAH to AIS, MIA and ADC (Hu et al., 2019) suggesting that early carcinogenesis of lung ADC is a gradual process shaped by continuous interaction with host immune surveillance. Importantly, dynamic immune activation and suppression have already started at preneoplastic stage. This is in line with a similar study on lung squamous cell carcinoma (SCC) precursors by Mascaux and colleagues, which revealed evidence of both immune activation and evasion along with evolution of preinvasive SCC lesions (Mascaux et al., 2019). Taken together, these data advocate for therapy targeting the immune microenvironment in patients with lung cancer precursors to prevent development of invasive lung cancers.

Identification of the molecular basis underlying immune activation and evasion may provide novel insights for understanding tumor-immune interactions and establishing biomarkers to select patients who may benefit from immunoprevention. WES and RRBS data available from the same cohort of pulmonary nodules provided the opportunity to depict the genomic and epigenetic features associated with the distinct immune landscape (Hu et al., 2020; Hu et al., 2019). Overall, oncogene mutations, HLA loss, CNV burden, AI burden and methylation status were all found to associate with the immune contexture, similarly to advanced cancers (Fridman et al., 2017; Thorsson et al., 2018). For example, *KRAS* mutations correlated with more active immune response, while *EGFR* mutations, HLA loss, higher CNV burden and decreased global methylation status correlated with a cold immune microenvironment. However, associations with any single genomic or epigenetic feature were weak, suggesting heterogenous but convergent evolution towards immune escape during early lung carcinogenesis.

Our previous work suggested that early lung ADC carcinogenesis may predominantly follow the clonal sweep model, whereby certain subclones in early-stage preneoplasia turn into dominant clones in later-stage diseases while unfit subclones are eliminated (Hu et al., 2019), primarily by the host immune system. In the early phases of carcinogenesis, stochastic genomic and epigenetic alterations lead to heterogeneous subclones with various combinations of molecular features that define the distinct biology of each subclone including survival ability under selective immune pressure. The impact of genomic and epigenetic alterations on immune response is intertwined. For example, methylation may directly affect immune response by regulating expression of immune genes (Liu et al., 2017) or potential neoantigens (Serrano et al., 2011) or indirectly by increasing DNA vulnerability for development of CNV and somatic mutations that can subsequently influence the immune microenvironment (Bakhoum and Cantley, 2018; Porta-Pardo and Godzik, 2016). Global hypomethylation is known to associate with CIN (Eden et al., 2003) and increased rate of somatic mutations (Chen et al., 1998) and indeed, global methylation level was negatively associated with TMB and CNV burden in this cohort of lesions (Hu et al., 2020). Since high CNV burden is associated with a cold immune microenvironment (Davoli et al., 2017) while high TMB can increase tumor immunogenicity and facilitate immune recognition and elimination of cancer cells (Jung et al., 2019), the impact of global hypomethylation (associated with both high CNV burden and TMB) on anti-cancer immunity is further complicated. In the end, the outgrowth of subclones is determined by the accumulated effects of all molecular aberrations. Only the cells with the ideal combination of molecular features enabling their rapid proliferation and escape from immune attack will survive and outgrow into the dominant clones in invasive cancers. For example, though invasive lung ADCs tend to harbor a high mutation burden that could lead to more active anti-tumor immune response, they also exhibited a higher CNV burden, higher likelihood of losing HLA and decreased global methylation, all of which are associated with a cold tumor immune microenvironment (Jeschke et al., 2017; Neal et al., 2018; Paulson et al., 2018; Rosenthal et al., 2019), resulting in an overall cold immune microenvironment in invasive ADCs (lower CD8+ T cell infiltration, CTLs, and T cell clonality, with higher CD4+ T cell infiltration and Tregs).

With the increasing implementation of CT-guided screening and advent of high-resolution diagnostic CT scans, there has been a drastic increase in the detection of IPNs (Bellomi et al., 2010; McWilliams et al., 2013), many of which are lung ADC precursors. However, their management remains controversial. Although surgical resection could potentially offer cure in a large proportion of these patients, surgery-associated morbidity and high cost have called surgical resection into question. Additionally, ~25% patients often present with multifocal diseases, which complicates surgery. We have previously demonstrated (Hu et al., 2019; Sivakumar et al., 2017) that preinvasive lung ADC precursors were molecularly simpler, and therefore theoretically easier to eradicate. In the current study, we showed that lung ADC precursors may exhibit an overall better-preserved anti-tumor immune landscape. In light of these findings, therapeutic strategies reprograming the tumor immune microenvironment in patients with lung ADC precursors prior to further immunosuppression in invasive lung cancers may be beneficial. The lung cancer immunoprevention clinical trial IMPRINT-Lung (NCT03634241) recruiting individuals with high-risk IPNs is currently underway to validate this hypothesis.

## Data Availability

All data may be found within the main manuscript or supplementary Information or available from the authors upon request.

## ACKNOWLEDGEMENTS

This study was supported by the MD Anderson Khalifa Scholar Award, the National Cancer Institute of the National Institute of Health Research Project Grant (R01CA234629-01), the AACR-Johnson & Johnson Lung Cancer Innovation Science Grant (18-90-52-ZHAN), the MD Anderson Physician Scientist Program, the MD Anderson Lung Cancer Moon Shot Program, Sabin Family Foundation Award, Duncan Family Institute Cancer Prevention Research Seed Funding Program, the Cancer Prevention and Research Institute of Texas Multi-Investigator Research Award grant (RP160668) and the UT Lung Specialized Programs of Research Excellence Grant (P50CA70907), Cancer Prevention and Research Institute of Texas (CPRIT) grant RP150079. We thank Sally Boyd, Jinzhen Chen, and Rong Yao, Eric Sisson and Stan Bujnowski for providing excellent technical support for high-performance cluster resource http://hpcweb.mdanderson.edu/citing.html. We thank MD Anderson Cancer Center’s Epigenomics Profiling Core and its Science Park Next-Generation Sequencing Core (supported by CPRIT Core Facility Support Award #RP 120348) for performing RRBS profiling.

## AUTHOR CONTRIBUTIONS

J.Z., I.W., H.K., and A.R. conceived, designed, and directed the study. H.D., X.H., R.C., J.F., L.L., C.G., C.C., and J.Y. performed the experiments. X.H., Jiexin Z., N.M., S.H., J.L., and Jianhua Z. conducted bioinformatic analyses. J.F., E.P., C.H., D.D., L.S., D.S., J.F., H.P., B.Z., L.Y., performed pathologic assessment and tissue processing. M.E., M.G., M.A., B.S., H.P., C.B., A.V., J.H., P.S., and P.A.F participated in data interpretation and clinical correlation. J.Z., R.C., H.D., H.K., A.R., and X.H. wrote the manuscript with comments from all authors.

## STAR METHODS

### Patients and tissue processing

Specimens were collected from 53 patients presenting with pulmonary nodules, who underwent surgical resection at Nagasaki Hospital (Japan) or Zhejiang Cancer Hospital (China) from 2014 to 2017 as described previously (Hu et al., 2019). The study was approved by the Institutional Review Boards (IRB) at MD Anderson Cancer Center, Nagasaki University Graduate School of Biomedical Sciences and Zhejiang Cancer Hospital. Hematoxylin and eosin (HE) slides of each case were reviewed by experienced lung cancer pathologists to confirm the diagnosis before further analyses.

### DNA and RNA extraction

After pathologic assessment, manual macro-dissection was performed to enrich premalignant cells or cancer cells for DNA or RNA extraction. DNA or RNA was isolated using the AllPrep® DNA/RNA FFPE Kit (Qiagen, Hilden, Germany) according to manufacturer’s instructions. Finally, the DNA samples were quantified by NanoDrop 1000 Spectrophotometer (Thermo Scientific, Wilmington, DE, USA) and RNA was quantified using the RNA High sensitivity kit on the Qubit 3.0 fluorometer (Thermo Fisher Scientific, USA). RNA quality and integrity were evaluated with RNA integrity number (RIN) (Schroeder et al., 2006), concentration (ng/μl), and size (nt) using RNA Screen Tape in 4200 Tape Station System (Agilent Technologies, USA).

### Gene profiling of immune cells using nCounter® platform

The nCounter® PanCancer Immune Profiling Panel (NanoString Technologies, Inc., Seattle, WA, USA), which contains 770 genes (including 730 immune associated genes and 40 housekeeping genes) using NanoString nCounter Analysis was applied for gene expression profiling as previously described (Cesano, 2015; Kulkarni, 2011). The data were imported into the nSolver 4.0 software (NanoString Technologies) for quality control (QC). After the QC was performed with default setting, background correction was done with negative controls, and data was normalized by using the geometric mean of the 6 positive controls, and 40 housekeeping genes. We further performed quantile-normalization and log2 transformation to stabilize the variance. A one-way ANOVA test was applied to identify differentially expressed genes (DEGs) in different stages. We modeled the p-values using a beta-uniform mixture (BUM) model, combined with false discovery rate (FDR) to determine a cutoff for p-values (Pounds and Morris, 2003). The DEGs, along with log ratios were then evaluated with Ingenuity Pathway Analysis (IPA) software (Quiagen, Hilden, Germany) (Kramer et al., 2014) to identify pathways that are enriched by these DEGs. We performed a Core Analysis with species set to human and tissue set to lung. IPA identifies the top canonical pathways associated with the list of DEGs by the Fisher’s exact test to ascertain enrichment. IPA also calculates a z-score to predict activation status of the pathway by comparing input genes and the stored activity pattern.

### Multiplex immunofluorescence staining

All samples were confirmed to be appropriate for multiplex immunofluorescence (mIF) analysis based on tissue quality and availability using the HE staining of same lesion by the experienced pathologist. The manual mIF staining was performed on unstained slides of FFPE samples using the Opal 7-Color fIHC Kit (Akoya Biosciences, USA) as previously described (Parra et al., 2018). Eight immune markers were placed in two 6-antibody panels before the stained slides were scanned by Vectra multispectral microscope (Akoya Biosciences, USA). Panel 1 contained pancytokeratin (AE1/AE3; epithelial marker; dilution 1:300; Dako, Carpinteria, CA), PD-L1 (clone E1L3N, dilution 1:100; Cell Signaling Technology, Beverly, MA), PD1 (clone EPR4877-2, dilution 1:250; Abcam, Cambridge, MA), CD3 (T lymphocyte marker; dilution 1:100; Dako), CD8 (cytotoxic T cell marker; clone C8/144B, dilution 1:20; Thermo Fisher Scientific, Waltham, MA), and CD68 (macrophage marker; clone PG-M1, dilution 1:450; Dako). Panel 2 contained pancytokeratin, CD3, CD8, CD45RO (memory T cell marker; clone UCHL1, ready to use; Leica Biosystems, Buffalo Grove, IL), Granzyme B (cytotoxic lymphocyte marker; clone F1, ready to use; Leica Biosystems), and FoxP3 (regulatory T cell marker; clone 206D, dilution 1:50; BioLegend, SanDiego, CA). Human tonsil FFPE tissues were used with each individual antibody on same fluorochrome assay to build the spectral library, and were also used with and without primary antibodies as positive and negative (autofluorescence) controls, respectively. The stained slides were scanned with Vectra 3.0 multispectral microscope system (Akoya Biosciences, USA) under fluorescent illumination. These mIF assays followed our previous publication (Francisco-Cruz et al., 2020).

### Multispectral analysis

The five individual fields for multispectral analysis were selected from areas of interest in a scanned low magnification (×10) image on Phenochart1.0.9 (Akoya Biosciences, USA). The five high magnification (×20) fields for mIF analysis were carefully selected by experienced pathologists after comparing with HE slides to capture malignant and premalignant cell cluster and various elements of heterogeneity. The corresponding normal fields were selected in the farthest field of tumor periphery with morphological normal tissue on the same slide (**Figure S5A**). The selected high magnification field was total 1.6725 mm^2^ in size (669×500μm per field) (**Figure S5B**). Each field with panel 1 and panel 2 were overlapped with sequential sections. The target areas were analyzed by in Form 2.4.4 software (Akoya Biosciences, USA). The area was divided into two compartments: the epithelial compartment (the alveolar wall and septal or malignant cell nests) and alveolar air space or tumor stroma compartment (**Figure S5C**). The individual cells were recognized by DAPI nuclei staining (**Figure S5D**). Panel 1 contained the co-localization patterns as follows: PD-L1 expressing CK+ cells (CK+PD-L1+), total T lymphocytes (total CD3+), cytotoxic T lymphocytes (CD3+CD8+), antigen experienced T lymphocytes (CD3+PD-1), antigen experienced cytotoxic T lymphocytes (CD3+CD8+PD- 1+), total macrophages (total CD68+), and PD-L1 expressing macrophages (CD68+ PD- L1+). Panel 2 contained the co-localization patterns as follows: total T lymphocytes (total CD3+), cytotoxic T lymphocytes (CD3+CD8+), activated cytotoxic T lymphocytes (CD3+CD8+GB+), memory T lymphocytes (CD3+CD45RO+), effector/memory T lymphocytes (CD3+CD8+CD45RO+), regulatory T lymphocytes (CD3+FOXP3+CD8-), and memory/regulatory T lymphocytes (CD3+CD45RO+FOXP3+) (**Figure S5**). The percentage of the total nucleated cell density from each individual cell phenotyping population was used for further analysis.

### TCRβ amplification and sequencing

Immunosequencing of the CDR3 regions of human TCRβ chains was performed using the protocol of ImmunoSeq (Adaptive Biotechnologies, hsTCRβ Kit) and T cell clonality and diversity were calculated as described previously (Reuben et al., 2017; Reuben et al., 2020).

Briefly, T cell clonality is a metric of T cell proliferation and reactivity, and it is defined as 1-Pielou’s evenness and is calculated on productive rearrangements by:

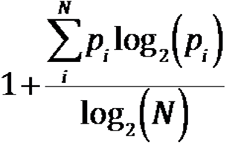

Where p_i_ is the proportional abundance of rearrangement i, and N is the total number of rearrangements. Clonality ranges from 0 to 1: values approaching 0 indicate a very even distribution frequency of different clones (polyclonal), whereas values approaching 1 indicate a distinct asymmetric distribution in which a few activated clones are present at high frequencies (monoclonal).

We also applied Inverse Simpson in order to observe T cell diversity, the sum over all observed rearrangements of the square fractional abundances of each rearrangements using productive templates.

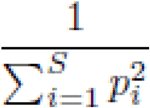

Where p_i_ is the proportional abundance of rearrangement i, and S is the total number of rearrangements. Inverse Simpson ranges from 1 to infinite, where a sample with little variation or abundance has a value approaching 1, and a maximally diverse and evenly distributed sample has a value approaching infinite.

### Analysis of genomic and methylation profiling data

Genomic and methylation data were generated and processed in previous studies as described (Hu et al., 2020; Hu et al., 2019). Somatic mutations, allelic imbalance (AI), copy number variations (CNV), oncogene mutations, global methylation data were directed from above mentioned studies. The following additional analyses were performed on the genomic data.

### Detection of allele-specific HLA loss

Class I HLA alleles for each HLA gene was inferred by POLYSOLVER using a two-step Bayesian classification approach (Shukla et al., 2015). This approach takes into account the base qualities of aligned reads, observed insert sizes, as well as the ethnicity-dependent prior probabilities of each allele (Shukla et al., 2015). Tumor purity and ploidy were estimated using ASCAT (Van Loo et al., 2010). We then applied LOHHLA (Loss Of Heterozygosity in Human Leukocyte Antigen) algorithm (McGranahan et al., 2017) to detect allele-specific HLA loss in each sample. In brief, logR and BAF across each HLA gene loci was obtained by binning the coverage at mismatch positions between homologous HLA alleles, and HLA haplotype specific copy numbers were then calculated based on logR and BAF value from the corresponding bin adjusted by tumor purity and ploidy. The median value of binned allelic copy number was used to determined LOH, where a copy number of < 0.5 indicated allele loss and AI was determined if p< 0.01.

### Statistical Analysis

Different statistical models were applied to assess the association among immune data, genomic data and methylation data. When assessing association between two variables, different tests were applied depending on the types of variables. For association between two continuous variables, spearman’s rank correlation test was used. For association between one continuous variable and one categorical variable, Wilcoxon rank-sum test (categorical variable with two levels) and Kruskal-Wallis test (categorical variable with more than two levels) were applied. The FDR method was used for multiple testing adjustment of p-values (Benjamini and Hochberg, 1995). All p-values are calculated with two-sided test, and p<0.05 was considered to be statistically significant.

## DATA AVAILABILITY

The data from WES has been deposited at European Genome-phenome Archive (EGA), which is hosted by The European Bioinformatics Institute (EBI) and the Centre for Genomic Regulation (CRG) under the accession code: EGAS00001004960 [https://www.ebi.ac.uk/ega/datasets/EGAD00001004960]. All other data may be found within the main manuscript or supplementary Information or available from the authors upon request.

## SUPPLEMENTAL INFORMATION

### SUPPLEMENTAL FIGURE LEGENDS

**Figure S1.**
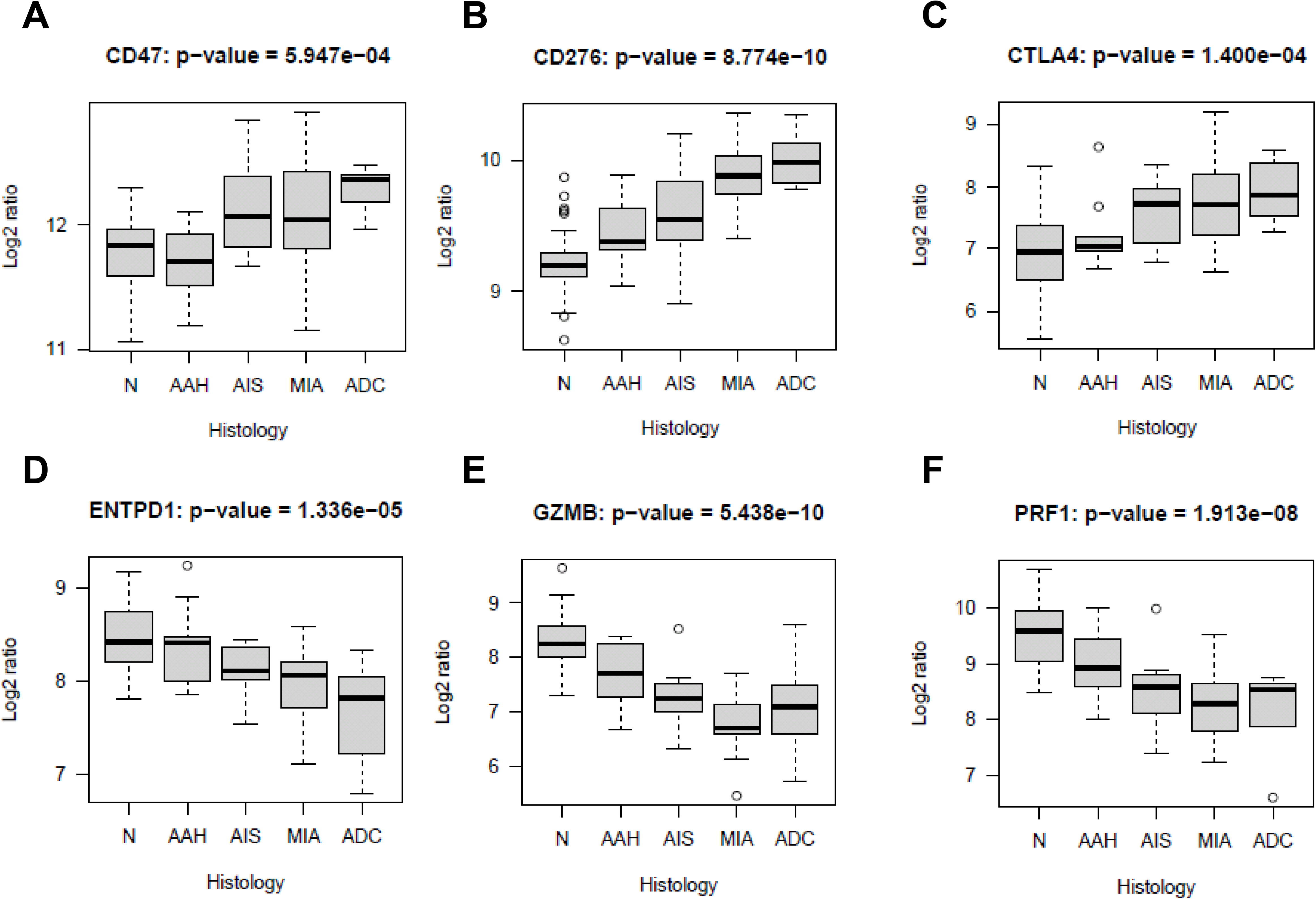
Example genes differentially expressed from preneoplasia to invasive lung adenocarcinoma. Gene expression of *CD47* (**A**), *CD276* (**B**), *CTLA4* (**C**), *ENTPD1* (**D**), *GZMB* (**E**), *PRF1* (**F**) was performed using the nCounter PanCancer Immune Profiling Panel (Nanostring). The difference in expression of each gene among different stages was evaluated using the Kruskal-Wallis H test. AAH: typical adenomatous hyperplasia. AIS: adenocarcinoma in situ. MIA: minimally invasive adenocarcinoma. ADC: invasive adenocarcinoma.

**Figure S2.**
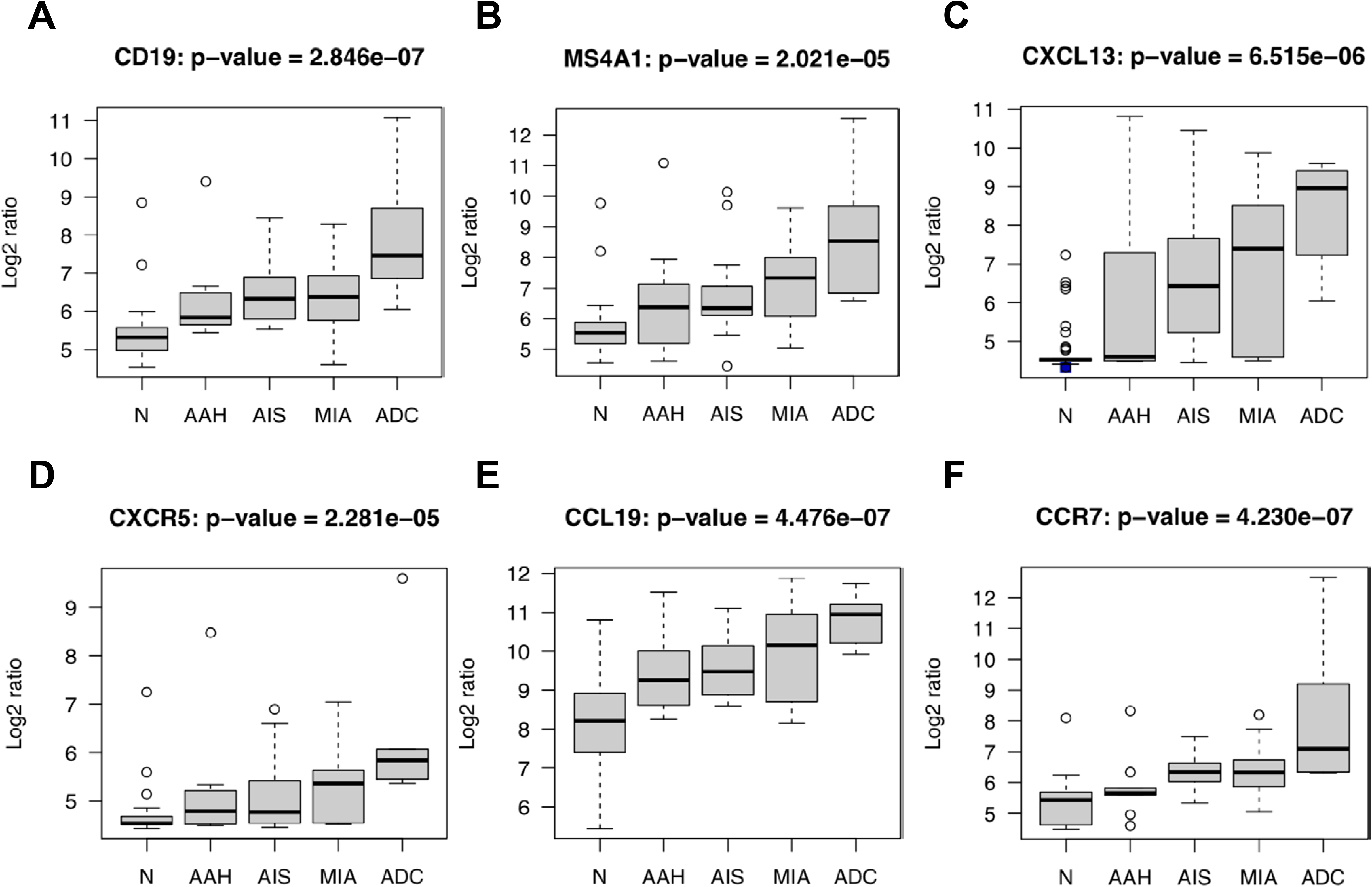
Expression of tertiary lymphoid structure (TLS) marker genes from preneoplasia to invasive lung adenocarcinoma. Gene expression of B cell marker *CD19* (**A**) and *MS4A1 (CD20)* (**B**); follicle formation markers *CXCL13* (**C**) and *CXCR5* (**D**); T lymphocyte homing markers *CCL19* (**E**) and *CCR7* (**F**) was performed using the nCounter PanCancer Immune Profiling Panel (Nanostring). The difference in expression of each gene among different stages was evaluated using the Kruskal-Wallis H test. AAH: typical adenomatous hyperplasia. AIS: adenocarcinoma in situ. MIA: minimally invasive adenocarcinoma. ADC: invasive adenocarcinoma.

**Figure S3.**
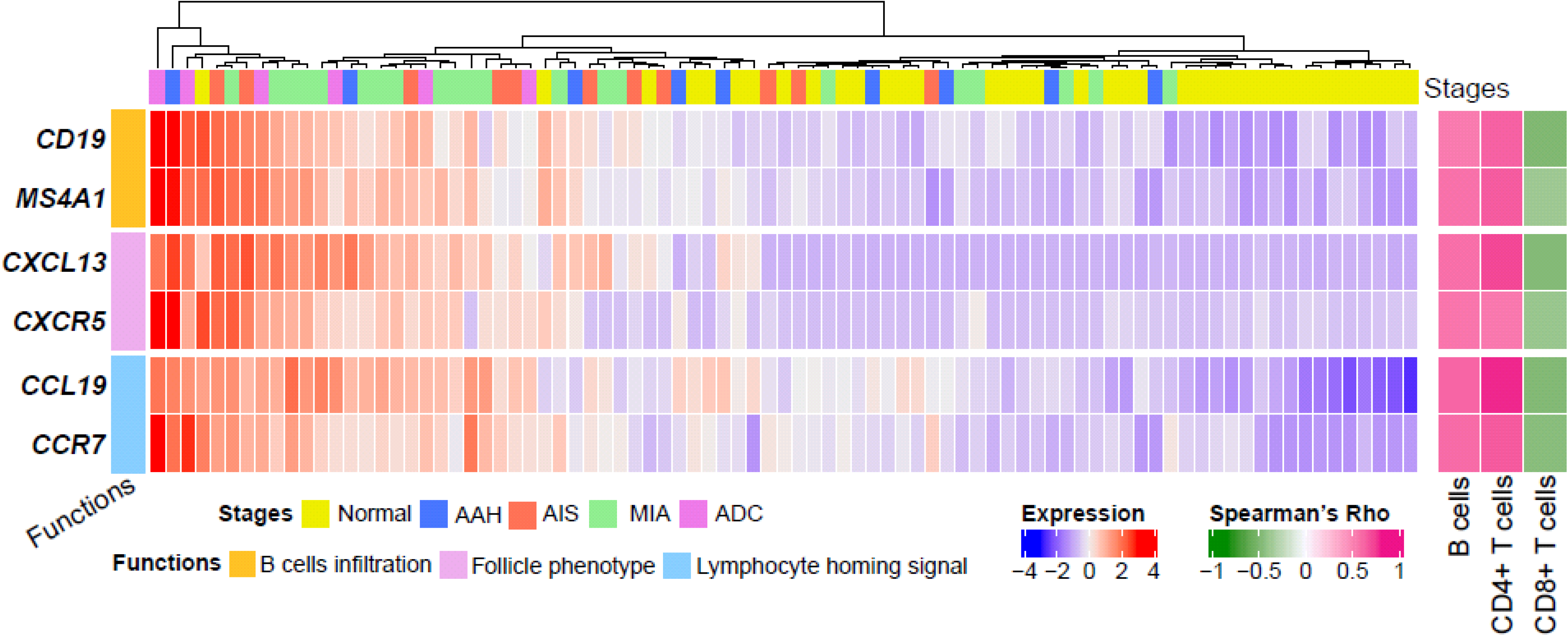
Expression of TLS marker genes was associated with B cells, CD4+ lymphocytes and CD8+ lymphocytes. Gene expression profiling was performed using the nCounter PanCancer Immune Profiling Panel (Nanostring). Infiltration of B cells, CD4+ T cells and CD8+ T cells was estimated using TIMER based on the gene expression using nCounter PanCancer Immune Profiling Panel. The association between the expression of each TLS marker gene and B cells, CD4+ T cells and CD8+ T cells were evaluated using xxx test.

**Figure S4.**
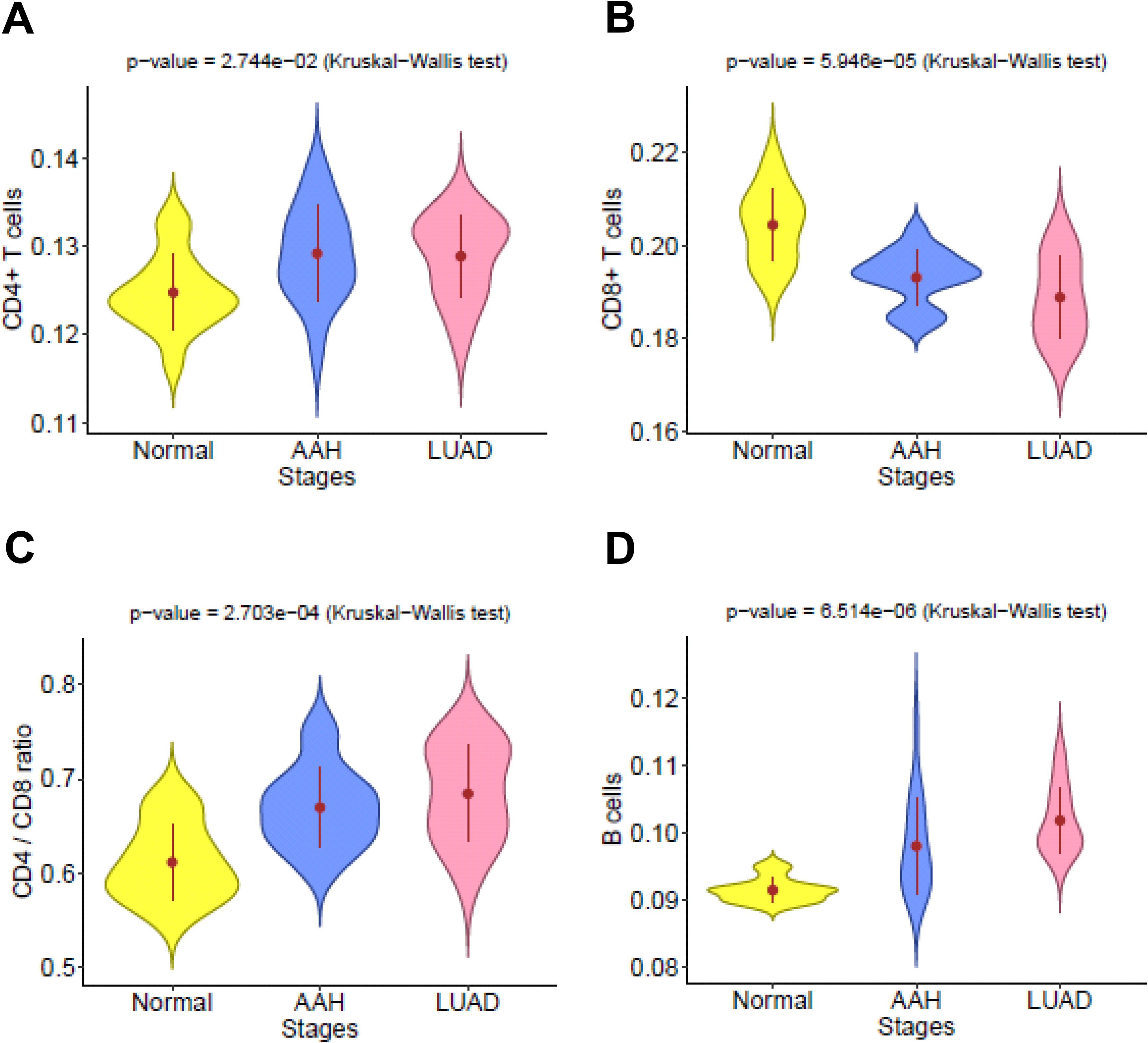
Progressively changes of immune cell in filtration normal lung, preneoplasia and invasive lung adenocarcinoma. Immune cell fractions including CD4+ T cells (A), CD8+ T cells (B), CD4 /CD8 ratio (**C**), B cells (**D**) were estimated using TIMER based on previously published RNA sequencing data from an independent cohort (GSE102511). Error bars indicate 95% confidence intervals and solid point represent mean value in each stage. The difference of cell fraction among stages was evaluated using the Kruskal-Wallis H test. NL: Normal lung tissue, AAH: Atypical adenomatous hyperplasia, ADC: Invasive adenocarcinoma.

**Figure S5.**
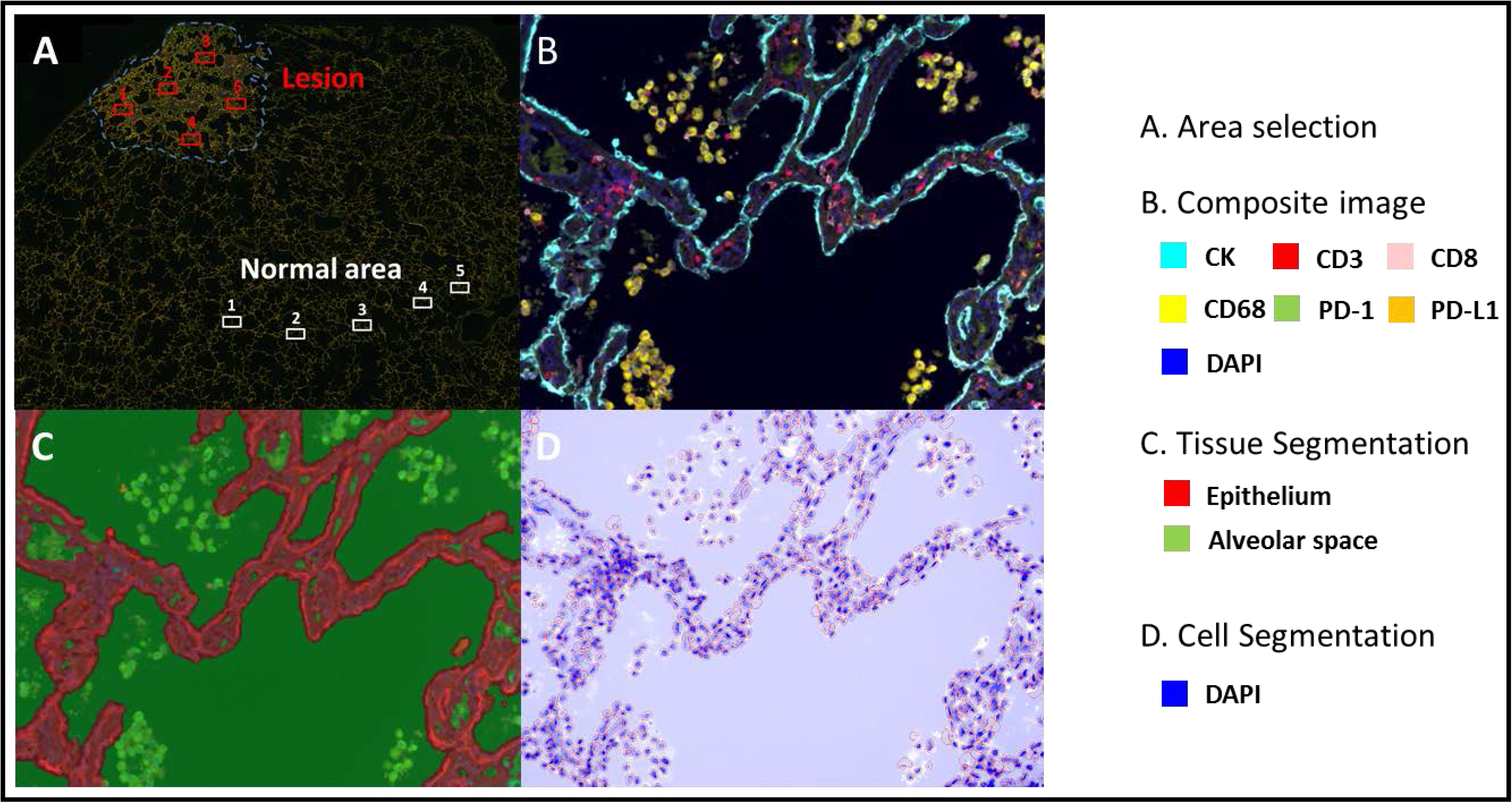
Quantitative image analysis of multiplex immunofluorescence image data. (**A**) After immunofluorescent staining, high magnification fields of interest were selected in both malignant (premalignant) and non-malignant regions. The non-malignant fields at the farthest regions from malignant (premalignant) regions with morphologically normal histology were selected. (**B**) Representative images from Panel 1 and Panel 2. (**C**) The selected fields were divided into two areas: epithelium and alveolar space. (**D**) Individual cells were recognized by DAPI (nuclear staining).

**Figure S6.**
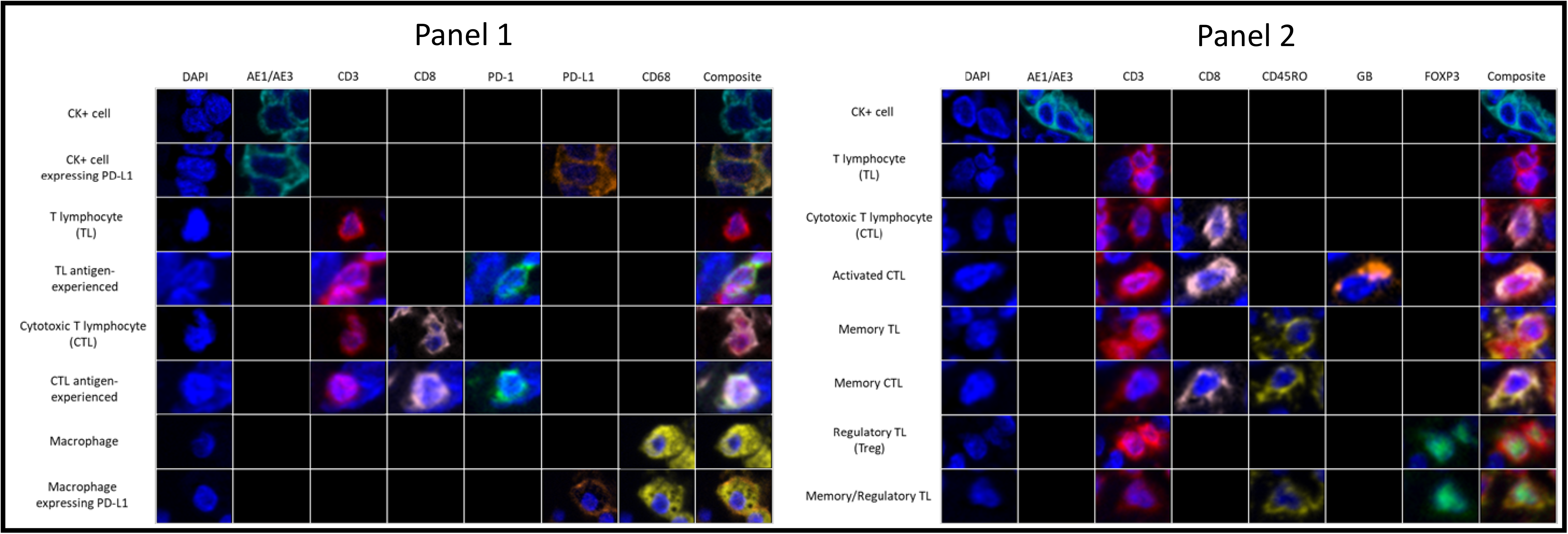
Subtyping immune and epithelial cells by multiplex immunofluorescence. Cell subtypes were defined as PD-L1 expressing epithelial cells (AE1/AE3+PD-L1+), T lymphocytes (CD3+), antigen-experienced T cells (CD3+PD-1+), cytotoxic T lymphocytes (CTL, CD3+CD8+), antigen-experienced CTL (CD3+CD8+PD- 1+), macrophages (CD68+) and PD-L1 expressing macrophages (CD68+PD-L1+) in panel 1 (left); and activated CTL (CD3+CD8+granzyme B+), memory T cell (CD3+CD45RO+), memory CTL (CD3+CD8+CD45RO+), regulatory T cell (CD3+CD8- FoxP3+), memory/regulatory T cell (CD3+CD45RO+FoxP3+) in panel 2 (right).

**Figure S7.**
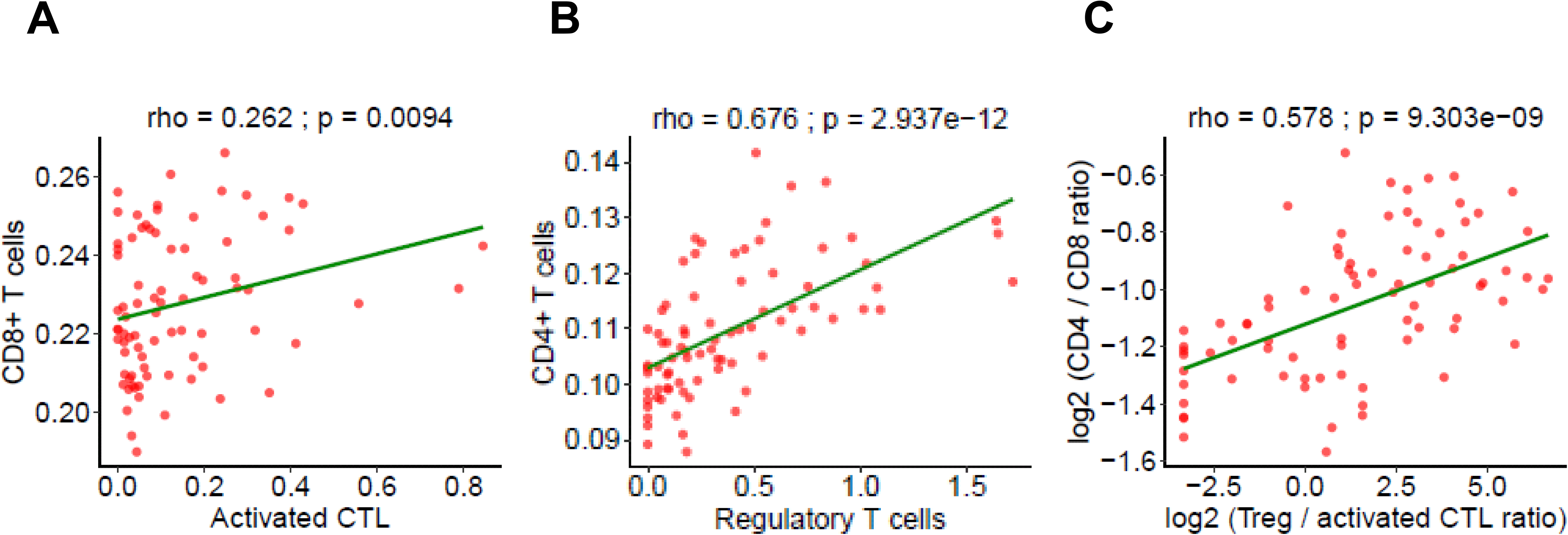
The correlation between immune cell subtypes measured by multiplex immunofluorescence and T cell subtypes inferred from immune gene expression. Fractions of immune cells including CD8+ T cells (**A**), CD4+ T cells (**B**) and CD4/CD8 ratio (**C**) estimated using TIMER based on the gene expression from nCounter PanCancer Immune Profiling Panel (y-axis) were correlated to activated CTL (CD3+CD8+granzyme B+), regulatory T cell (CD3+CD8-FoxP3+) and Treg/activated CTL ratio respectively (x-axis). All fraction and ratio were log2 transformed for visualization. The correction coefficient (rho) was assessed by Spearman’s rank correlation test.

**Figure S8.**
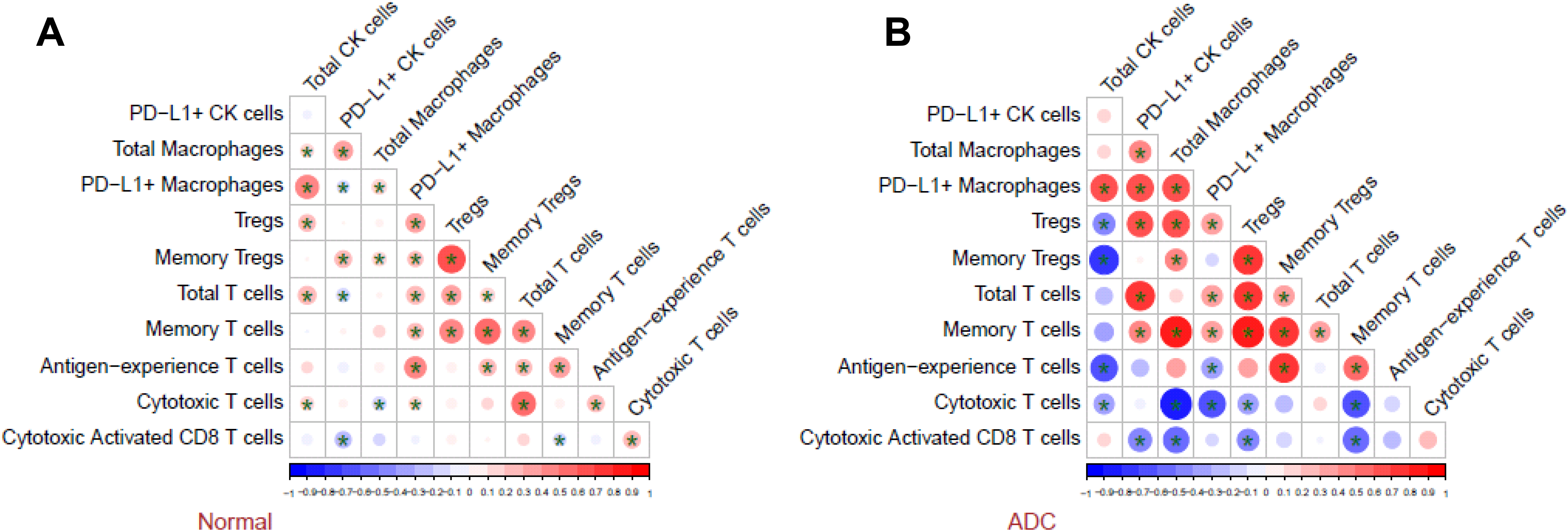
Various subtypes immune cells and their interaction with epithelial cells in normal lung tissues and invasive lung adenocarcinoma. The correlation between immune cell subtypes and CK+ epithelial cells measured by mIF in normal lung (**A**) and invasive lung adenocarcinoma (**B**). Significant correlation was marked with ^*^ (p<0.05). Red circles indicate positive correlations and blue circles indicate negative correlations. The size of circles is proportional to the spearman’s correlation co-efficient between each pair of cells. ADC: Invasive adenocarcinoma.

**Figure S9.**
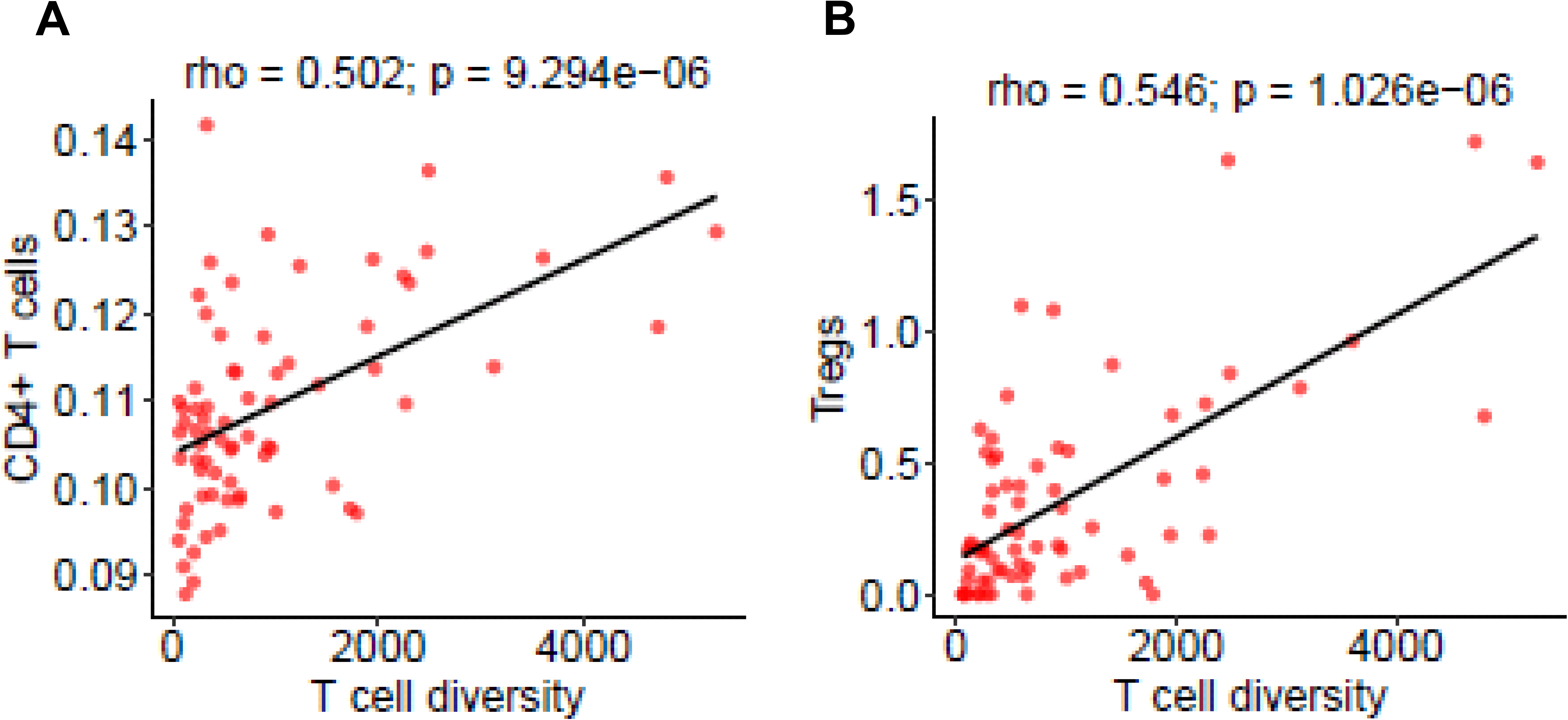
Correlation between T cell diversity and regulatory T cells. T cell diversity was positively correlated with infiltration of CD4+ T cells derived from immune gene expression using TIMER (**A**), regulatory T cells from multiplex immunofluorescence (mIF) (**B**). The correction coefficient (rho) was assessed by Spearman’s rank correlation test.

**Figure S10.**
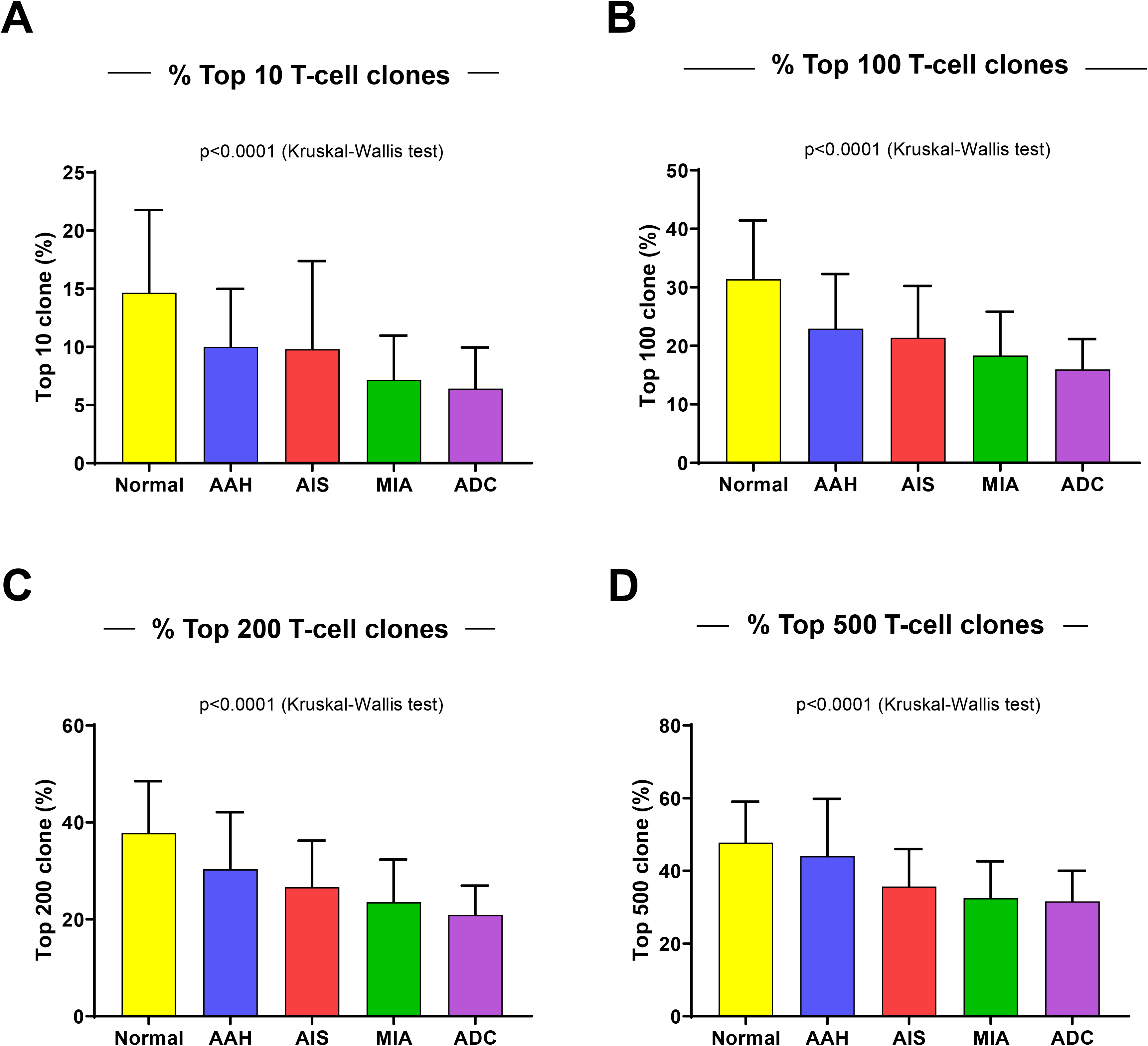
Progressive decrease of top T cell clone frequencies from preneoplasia to invasive lung adenocarcinoma. Frequencies of (**A**) top 10, (**B**) tope 100, (**C**) top 200 and (**D**) top 500 T cell clones in the Normal (yellow), AAH (blue), AIS (red), MIA (green) and ADC (purple). The difference between different stages were assessed by the Kruskal-Wallis H test.

**Figure S11.**
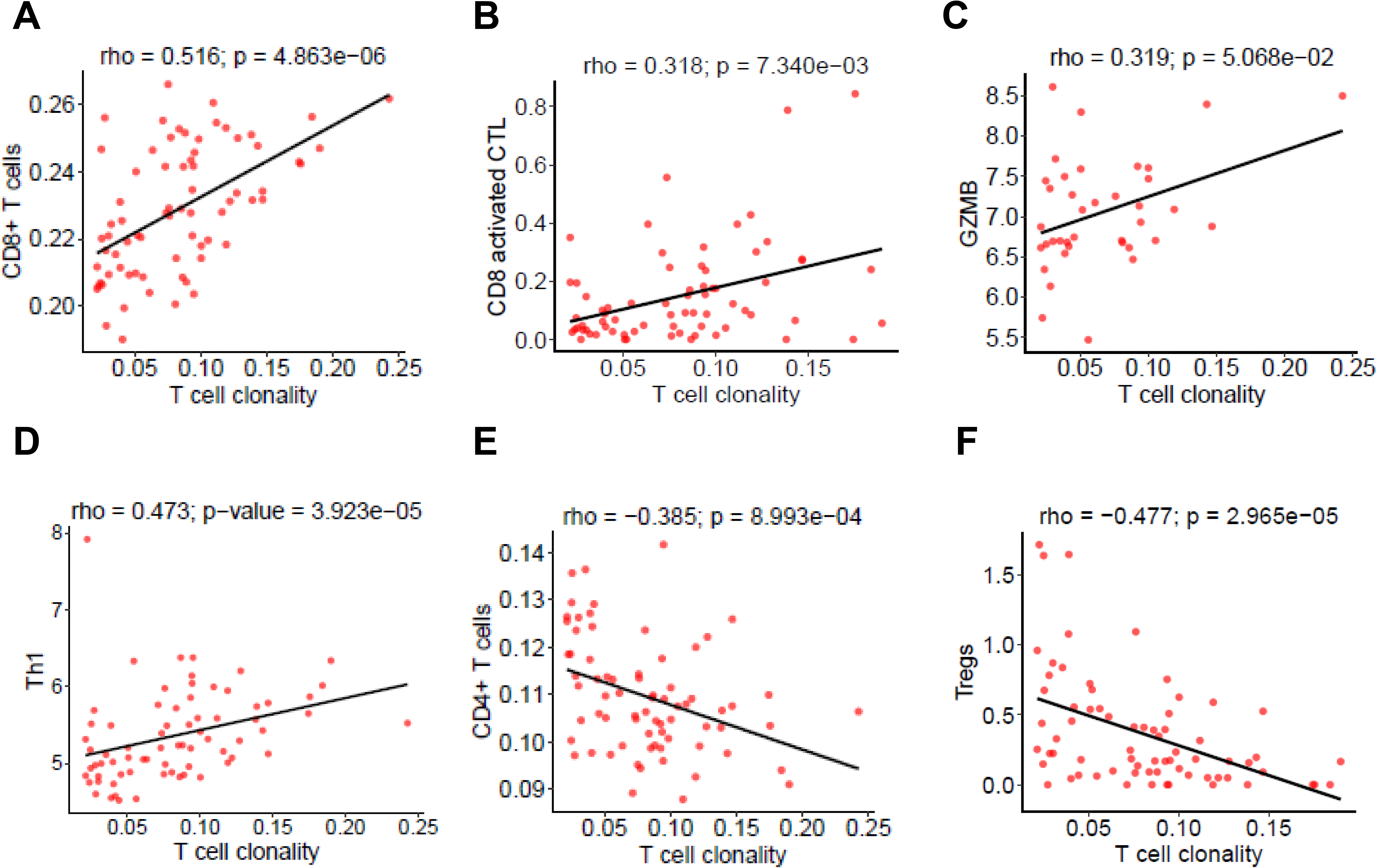
Correlation between T cell clonality and different T cell subtypes. T cell clonality was positively correlated with infiltration of CD8+ T cells inferred from gene expression by TIMER (**A**), activated cytotoxic T cells (**B**) by mIF, expression of GZBM (**C**) and Th1 cytokines (INFG, IL12A and IL12B) (**D**) but negative correlated with infiltration of CD4+ T cells derived from gene expression by TIMER (**E**) and regulatory T cells by mIF (**F**). The correction coefficient (rho) was assessed by Spearman’s rank correlation test.

**Figure S12.**
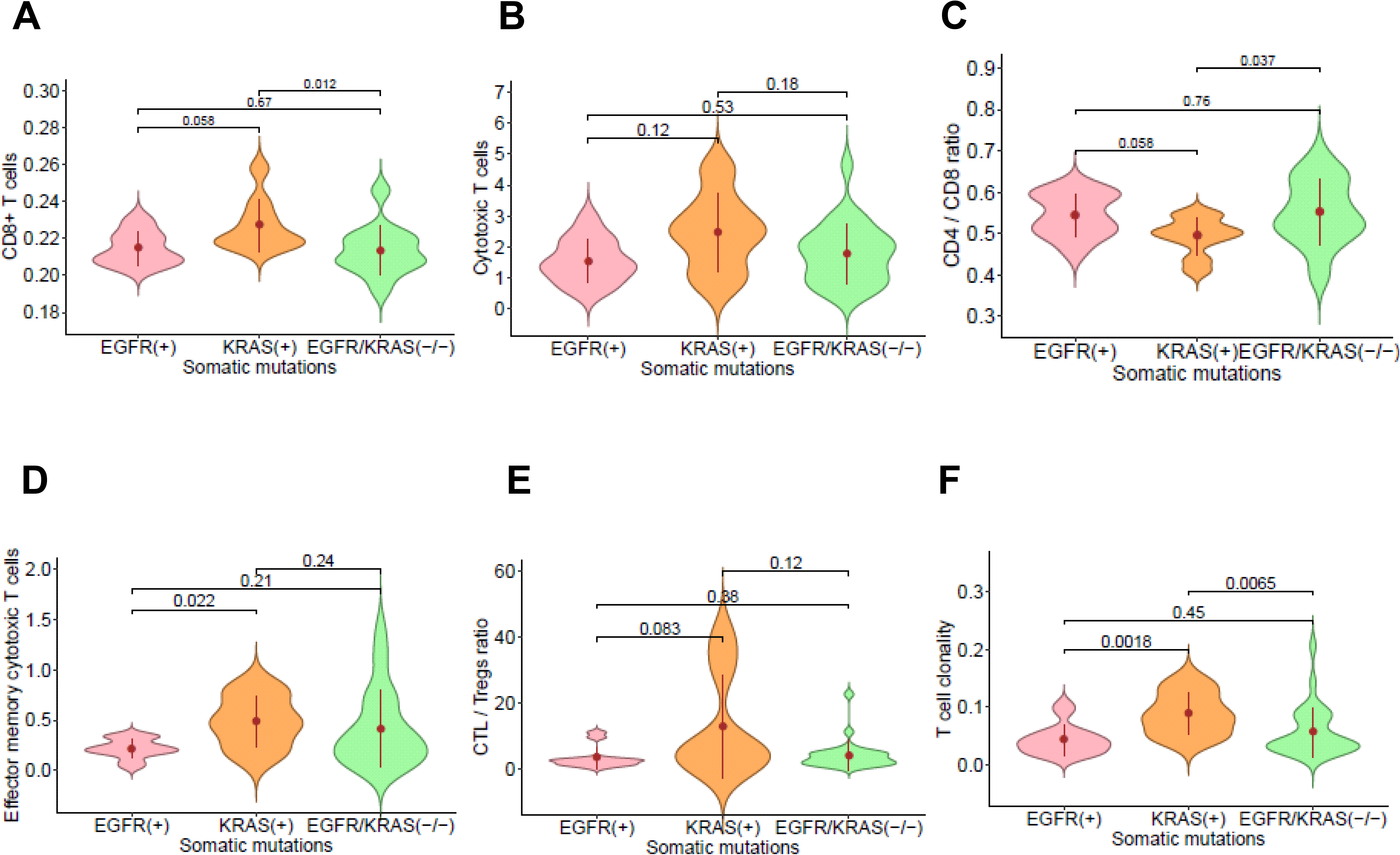
The impact of oncogene mutations on T cell features. Comparison of infiltration of CD8+ T cells inferred from gene expression by TIMER (**A**), cytotoxic T cells (CTL) (**B**) by mIF, CD4/CD8 ratio inferred from gene expression by TIMER (**C**), Effector memory cytotoxic T cells by mIF (**D**), CTL/Treg ratio (**E**) by mIF, T cell clonality by TCR sequencing (**F**) in lesions with *EGFR* mutation (pink), *KRAS* mutation (orange) and wildtype for both *KRAS* and *EGFR* (green). The difference was assessed by Wilcox-rank sum test.

**Figure S13.**
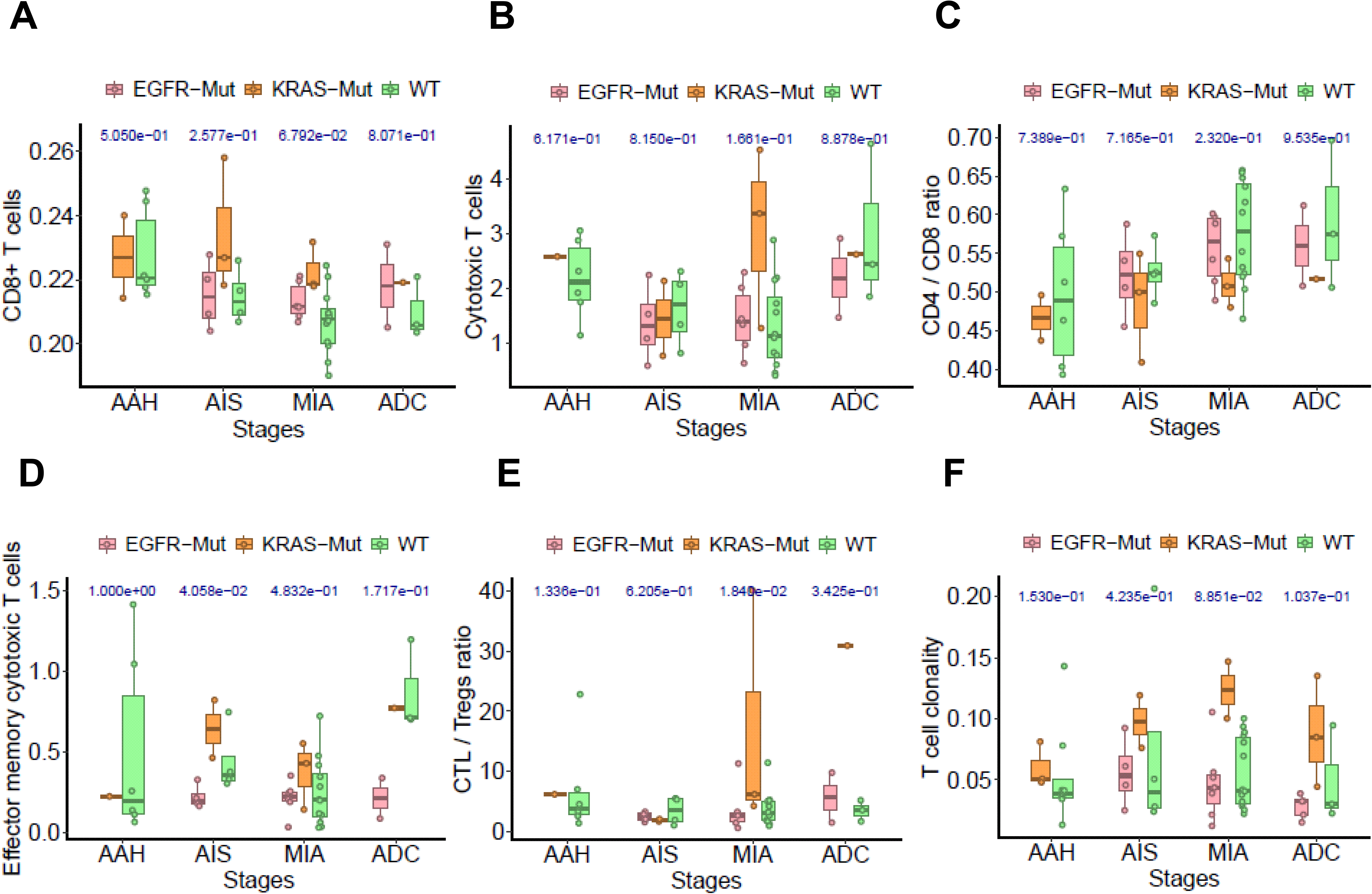
The impact of oncogene mutations on T cell features by different histologic stages. Comparison of infiltration of CD8+ T cells inferred from gene expression by TIMER (**A**), cytotoxic T cells (CTL) **B**) by mIF, CD4/CD8 ratio inferred from gene expression by TIMER (**C**), Effector memory cytotoxic T cells by mIF (**D**), CTL/Treg ratio (**E**) by mIF, T cell clonality by TCR sequencing (**F**) in lesions with *EGFR* mutation (pink), *KRAS* mutation (orange) and wildtype for both *KRAS* and *EGFR* (green) in AAH, AIS, MIA and ADC. The difference was assessed by Wilcox-rank sum test. AAH: Atypical adenomatous hyperplasia, AIS: Adenocarcinoma in situ, MIA: Minimally invasive adenocarcinoma, ADC: Invasive adenocarcinoma.

**Figure S14.**
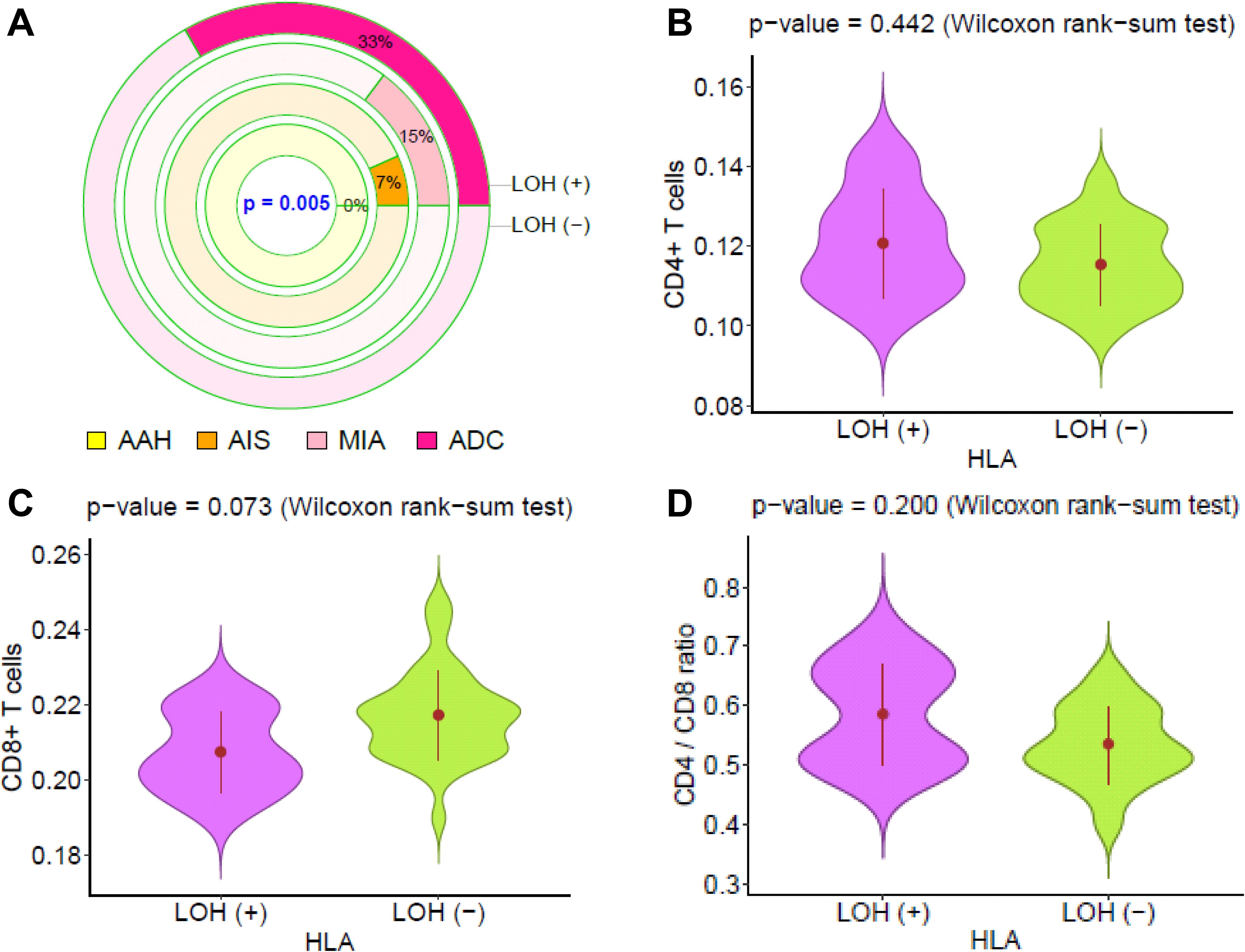
Loss of heterozygosity of HLA (HLA LOH) in different histologic stages from preneoplasia to invasive lung adenocarcinoma and its potential impact on immune contexture. (**A**) The proportion of AAH, AIS, MIA and ADC lesions had evidence of HLA-LOH. Chi-squared test were used to assess the difference among different histologic stages. The difference of infiltration of CD4+ T cells (**B**), CD8+ T cells (**C**) and CD4/CD8 ratio (**D**) inferred from gene expression by TIMER between lesions with (purple) and without (green) HLA-LOH.

**Figure S15.**
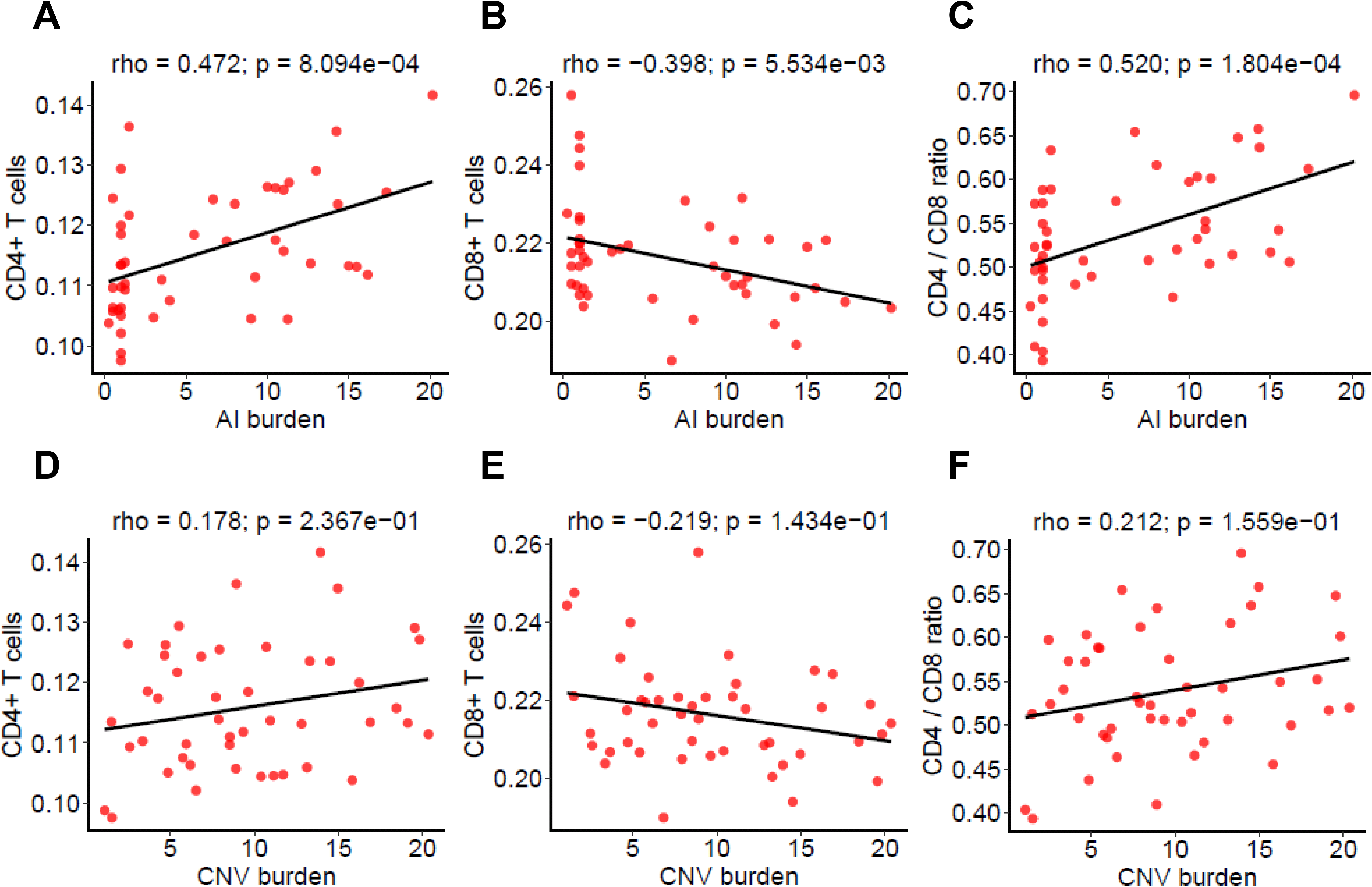
The potential impact of chromosomal copy number changes on immune infiltration. The correlation between allelic imbalance (AI) burden (number of AI events) and infiltration of CD4+ T cells (**A**), CD8+ T cells (**B**), CD4/CD8 ratio (**C**) inferred from gene expression by TIMER. The correlation between copy number variation (CNV) burden (normalized as the number of genes with CNV) and infiltration of CD4+ T cells (**D**), CD8+ T cells (**E**), CD4/CD8 ratio (**F**) inferred from gene expression by TIMER. The correction coefficient (rho) was assessed by Spearman’s rank correlation test.

**Figure S16.**
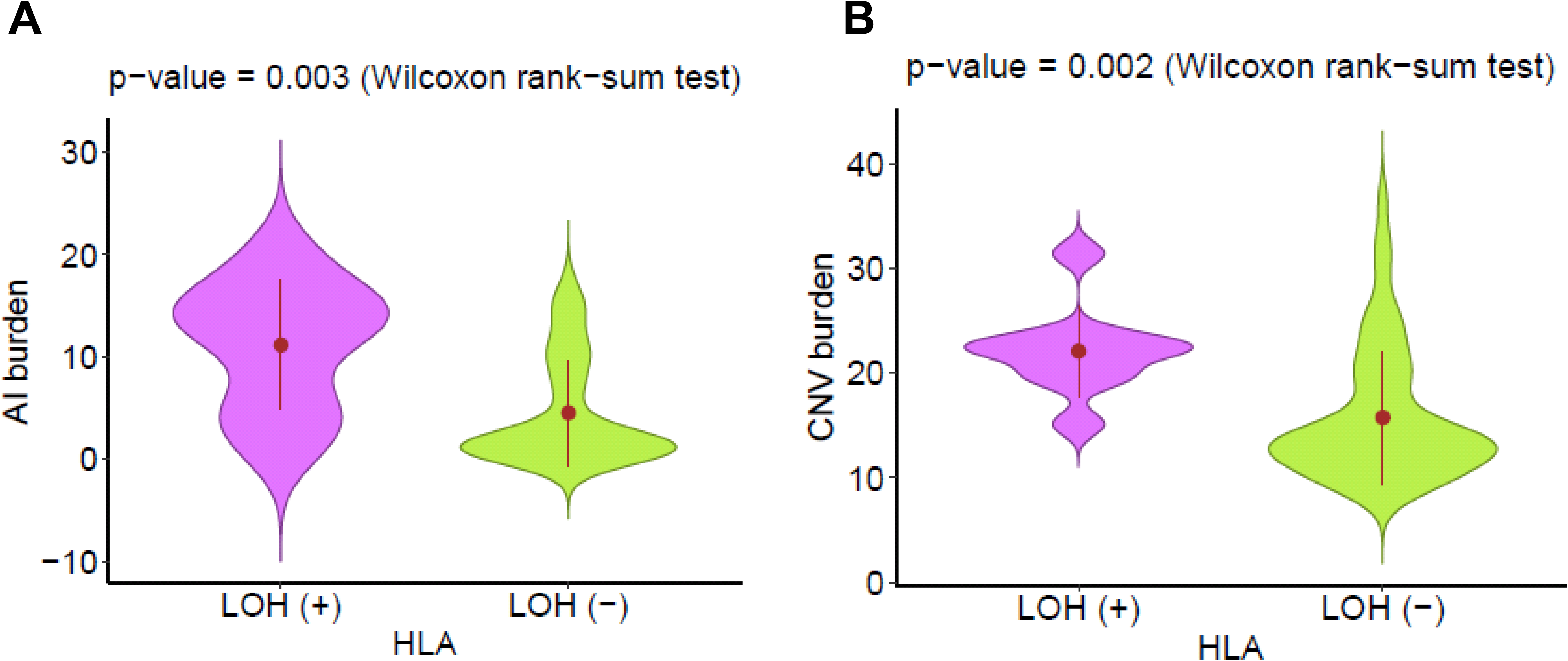
The relationship between HLA loss and chromosomal copy number variations. (**A**) Comparison of allelic imbalance (AI) burden (number of AI events) in lesions with (purple) and without (green) HLA-LOH. (**B**) Comparison of copy number variation (CNV) burden (normalized as the percent of genes with CNV) in lesions with (purple) and without (green) HLA-LOH. Willcoxon rank-sum test was used to assess the differences.

**Figure S17.**
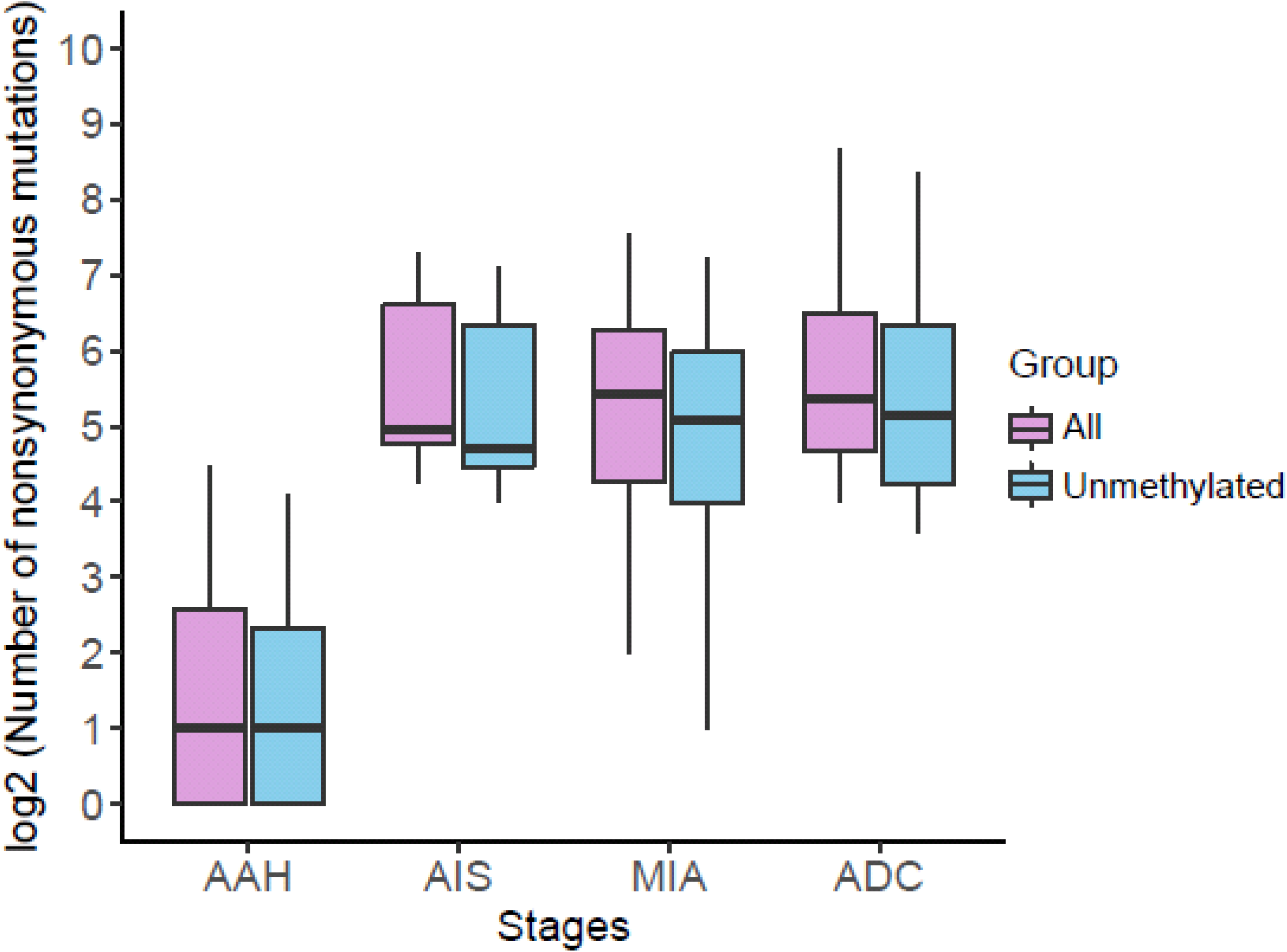
Promoter methylation and mutation burden from preneoplasia to invasive lung adenocarcinoma. The number of all nonsynonymous mutations in each histologic stage is shown as purple boxes and the number of nonsynonymous mutations from genes without promoter methylation (<30% CpG sites methylated) is shown as blue boxes.

**Figure S18.**
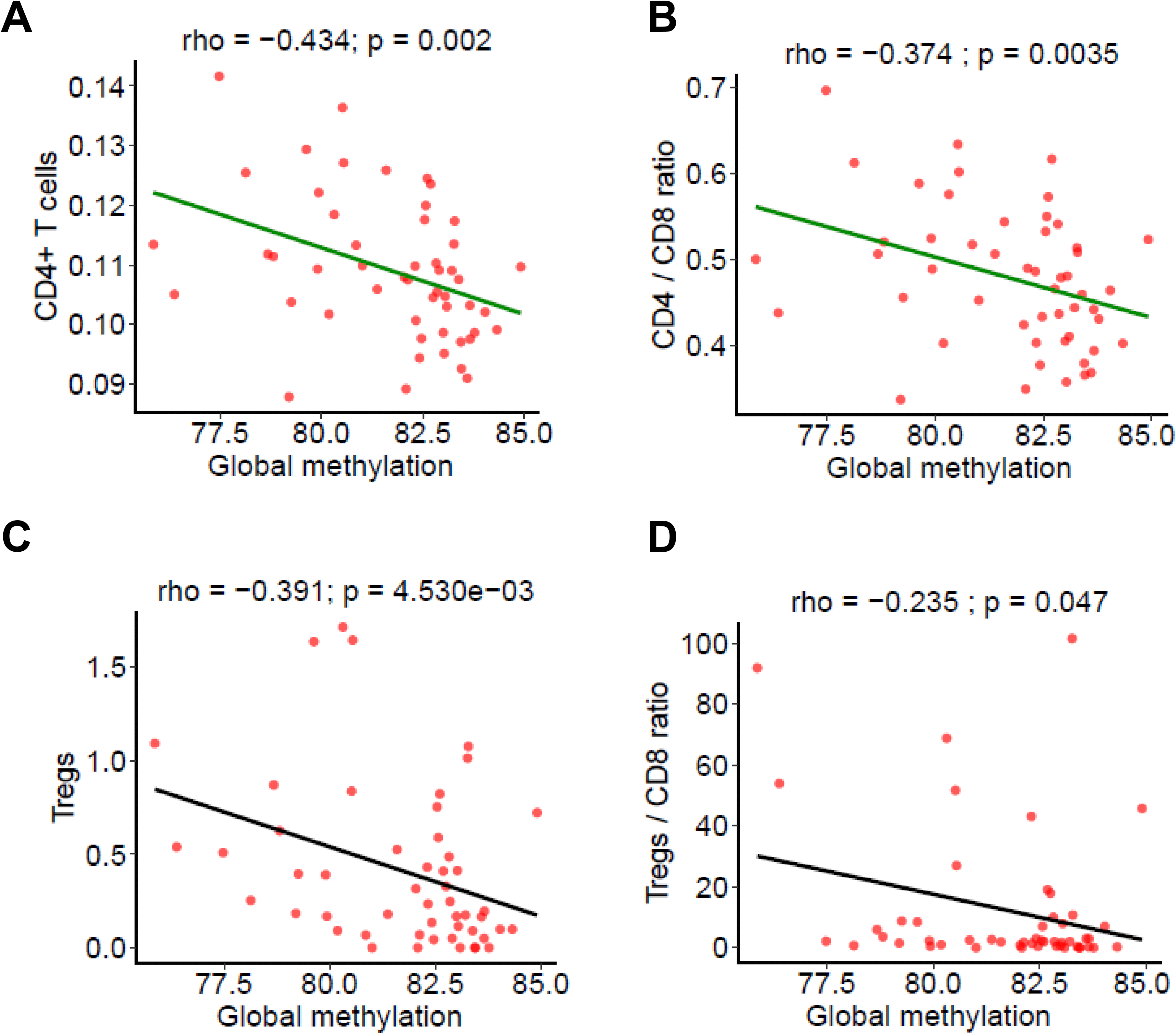
The potential impact of global methylation and immune infiltration. The correlation between global methylation levels (using LINE-1 as a surrogate marker) with CD4+ T cells (**A**), CD4/CD8 ratio (**B**) inferred from immune gene expression by TIMER as well as Tregs (**C**) and Treg/CD8 ratio (**D**) measured by mIF. The correction coefficient (rho) was assessed by Spearman’s rank correlation test.

### SUPPLEMENTAL TABLES

**Table S1. Clinical characteristics and availability of different immune and molecular data**

**Table S2. Statistics of Differentially Expressed Genes**

